# RNA-seq analysis of skeletal muscle in motor neurone disease cases and controls

**DOI:** 10.1101/2023.03.13.23287229

**Authors:** Anna Freydenzon, Shivangi Wani, Vanda Bharti, Leanne M. Wallace, Anjali K. Henders, Pamela A. McCombe, Robert D. Henderson, Frederik J. Steyn, Naomi R. Wray, Shyuan T. Ngo, Allan F. McRae

## Abstract

**Background:** Amyotrophic lateral sclerosis (ALS), the most predominant form of Motor Neuron Disease (MND), is a progressive and fatal neurodegenerative condition that spreads throughout the neuromotor system by afflicting upper and lower motor neurons. Lower motor neurons project from the central nervous system and innervate muscle fibres at motor endplates, which degrade over the course of the disease leading to muscle weakness. The direction of neurodegeration from or to the point of neuromuscular junctions and the role of muscle itself in pathogenesis has continued to be a topic of debate in ALS research.

**Methods:** To assess the variation in gene expression between affected and nonaffected muscle tissue that might lead to this local degeneration of motor units, we generated RNA-seq skeletal muscle transcriptomes from 28 MND cases and 18 healthy controls and conducted differential expression analyses on gene-level counts, as well as an isoform switching analysis on isoform-level counts.

**Results:** We identified 52 differentially-expressed genes (Benjamini-Hochberg-adjusted *p* < 0.05) within this comparison, including 38 protein coding, 9 long non-coding RNA, and 5 pseudogenes. Of protein-coding genes, 31 were upregulated in cases including with notable genes including the collagenic *COL25A1* (*p* = 3.1 × 10^−10^), *SAA1* which is released in response to tissue injury (*p* = 3.6 × 10^−5^) as well as others of the SAA family, and the actin-encoding *ACTC1* (*p* = 2.3 × 10^−5^). Additionally, we identified 17 genes which exhibited a functional isoform switch with likely functional consequences between cases and controls.

**Conclusions:** Our analyses provide evidence of increased tissue generation in MND cases, which likely serve to compensate for the degeneration of motor units and skeletal muscle.

## Background

Motor neuron disease (MND) is a group of several similar neurodegenerative disorders with highly variable patterns of onset, progression, and symptomology. Amyotrophic lateral sclerosis (ALS), the most predominant motor neuron disease, is identified as featuring both upper and lower motor neuron loss. There is active discussion as to whether MND operates as a ‘dying forward’ phenomenon where initial cell death occurs in the cortical motor neurons and extends down into the corticospinal projections (1), or a ‘dying back’ phenomenon where neuromuscular junctions (NMJs) are the first to suffer failure and the pathology proceeds up the axon back into the body of the motor neuron (2). Nevertheless, the progressive loss of upper and lower motor neurons results in the presentation of clinical features including spasticity and brisk reflexes, and muscle weakness and wasting across multiple body regions (3). In MND, muscle weakness and wasting occurs in response to the dismantling of neuromuscular junctions (NMJs) and axonal retraction that ultimately results in the denervation of skeletal muscle and subsequent die-off (4). Compensatory mechanisms can be seen in surviving motor units; axon terminals undergo sprouting to markedly increase reinnervation of skeletal muscle, and this occurs in parallel with abnormally fast firing (5).

MND is a highly polygenic disease consisting of over 20 known associated genes, although only ~10% of patients have familial history associated with mutations of large effect (4). One functional mechanism implicated by MND-associated genes is disturbance in RNA metabolism, which has contributed to an increasing research focus exploring transcriptomic machinery (6). One of the most notable genes is *TDP-43*, which undergoes substantial posttranslational modification and aggregation in both MND and frontotemporal dementia (7), leading to targeted RNA instability in its overexpression that results in consequences for ribosome biogenesis and oxidative phosphorylation pathways (8). Similarly, FUS is involved in transcriptional regulation through binding to RNA targets to regulate splicing and the removal of minor introns (9). Recent transcriptome-wide studies have been conducted on a variety of different tissues and models to find aberrant differential gene expression reflected in all tested tissue types, including but not limited to the postmortem motor cortex (10), patient whole blood (11), motor neurons from stem-cell derived spinal motor axons (12), and spinal cords from the recently generated PFN1 mouse model (13). Here, we report a whole transcriptome analysis of skeletal muscle tissue from MND patients, and describe the compensatory mechanisms invoked within MND-afflicted muscle tissue in response to denervation and local cell death.

## Methods

### Patient recruitment

Patient recruitment and muscle biopsies were undertaken as described by Steyn et al. 2020 (14). Briefly, MND patients who met the revised El-Escorial criteria for ALS (15) at the time of diagnosis were enrolled from the Royal Brisbane and Women’s Hospital MND clinic for the collection of skeletal muscle biopsies, along with healthy controls in the form of relatives, friends or community volunteers. For all participants, exclusion criteria were history of a metabolic condition (e.g. Hashimoto’s disease) and diabetes mellitus. Participants involved in the study are described in **Table S1**. Muscle biopsies (~200mg) were collected from the vastus lateralis from one leg (the least affected leg for MND participants) using a hollow Bergstrom biopsy needle modified for suction. Samples were placed in Dulbecco’s Modified EaglMedium/Ham’s F-12 with 0.5% gentamicin for transport to the laboratory. A portion (~50-100mg) of muscle tissue was divided from the primary biopsy, washed three times with room temperature phosphate buffered saline, frozen on dry ice, and stored at −80 degrees Celsius.

### RNA sequencing

RNA sequencing was performed on MND and healthy controls for both blood and muscle samples (36 overlapping; 23 cases, 13 controls). RNAseq was performed first on an initial batch with all blood and muscle samples, followed by the same muscle samples being re-run once (split across two additional batches) to increase total sequencing reads.

#### Muscle

Muscle samples passing QC were contributed by 17 healthy controls and 28 MND cases. Total RNA was extracted from the muscle tissue using a standard Trizol-chloroform extraction method. Post extraction, the total RNA was quantitated using the Epoch microplate spectrophotometer followed by quality assessment on the Agilent Bioanalyzer using the RNA Nano kit. On average the samples yielded 7.4 micrograms of total RNA with an average RNA Integrity Number (RIN) of 8.2.

One microgram of DNAse1 treated total RNA was then used to prepare libraries for sequencing. The libraries were prepared using Illumina’s Truseq stranded total RNA library prep gold kit in conjunction with the TruSeq RNA UD indexes. The libraries were then equimolar pooled and paired end sequencing was carried out on HiSeq 4000 system.

#### Blood

Blood samples passing QC were contributed to by 13 healthy controls and 24 MND cases. Approximately 2.5mL of whole blood was collected in PAXgene Blood RNA tubes. Total RNA was extracted from these tubes using the PAXgene Blood RNA kit from Qiagen. Post extraction, the total RNA was quantitated using the Nanodrop spectrophotometer followed by quality assessment on the Agilent Bioanalyzer using the RNA Nano kit. On average the samples yielded 4.8 micrograms of total RNA with an average RNA Integrity Number (RIN) of 8.2.

500ng of DNAse1 treated total RNA was then used to prepare libraries for sequencing. The libraries were prepared using Illumina Truseq stranded total RNA library prep gold kit in conjunction with the TruSeq RNA UD indexes. The libraries were then equimolar pooled and paired end sequencing was carried out on HiSeq 4000 system.

### Read processing

Initial QC and read trimming was undertaken using fastp (16), setting a threshold of 1 to pass through all non-empty reads (aberrant reads being filtered in the alignment step). By default, fastp conducts automatic adapter trimming by overlapping read pairs, as well as N-based qualify filtering and polyG tail trimming.

Trimming reads were mapped to the GENCODE v32-indexed genome through STAR 2.7.3 (17) with non-default parameters suggested by the STAR 2.7.0 manual for ENCODE alignment. These parameters specify that splices are only recognized if they fall over indexed, known splice junctions, sets a maximum number of mismatches to be read length* 0.04, limits genomic distance between read mates to 1,000,000 and sets a minimum intron length of 20bp.

The STAR-generated genome alignments were then quantified to gene meta-features through Rsubread featurecounts (18), with the parameter requiring both pairs to be mapped. Samples with alignment counts of < 20,000,000 across summed technical replicates were consequently filtered out of the analysis (4 blood, 1 muscle).

In addition, Salmon 1.0 (19) was run in quasi-mapping mode to the GENCODE v32 transcriptome (indexed over *k*-mer length 17). This was done to verify signals detected in STAR and act as a definitive source of transcripts as compared to the finer STAR genomic alignment method. This was run with the non-default parameters of validateMappings, seqBias, gcBias and useEM. This automatically estimates the library type (paired), runs in selective alignment mode where it ranks potential alignment sites based on mapping score, corrects for hexamer priming and GC biases and uses the expectation maximization (EM) method for abundance estimates.

Tximport 1.20 (20) was used to aggregate the Salmon transcript counts across samples to acquire both the summarized gene counts for the analysis complimentary to STAR, as well as the counts of the transcripts for differential isoform analyses.

### Differential expression analyses

STAR-generated read counts were read into DESeq2 1.30.1 (21) to collapse replicates into a single measure, and to estimate size factors and dispersion factors. Genes with ≥ 3 samples with normalized counts ≤ 5 were pruned from the individual blood and muscle sets. The resulting subset of genes were then run through DESeq2’s negative binomial GLM with a Wald significance test, increasing maximum iterations to 5,000.

The muscle and blood GLMs were fitted with MND as the condition and age and sex as covariates. Additionally, a binary variable indicating whether the samples were replicated across the second or third batch was added to the muscle model. These preliminary models were designated as the ‘base’ models.

The variation in counts was visualized within the muscle and blood samples through a principal component analysis applied to the 500 genes with highest variance across all samples, as well as through plotting the relative log expression on both centered and non-centered counts. Two control muscle samples were consequently dropped from further analysis due to featuring highly irregular counts and variance.

The genomic inflation factor (GIF) for each analysis (blood and muscle) was calculated as the median of the χ^2^ test statistic divided by the median of the χ^2^ distribution with one degree of freedom (22). The GIF is a summary statistic that compares the distribution of test statistics against expectation under the null hypothesis of no association of gene expression with MND and has an expected value of 1 when confounding is absent from the data, under the assumption that at least 50% of tests represent truly null associations. The GIF of the muscle models was found to be high, and as such a surrogate variable analysis (SVA) was conducted individually for blood and muscle using the *r* sva 3.38 package (23). SVA analysis estimates the probability of each gene being an empirical control gene (i.e., not associated with MND) through an iterative re-weighted least squares regression and captures variation outside of a specified model with known covariates. In this case, the SVA was conducted with MND status as the only covariate to preserve in the non-null model. This function also generated a suggested number of SVs to use in differential expression analysis through a permutation procedure.

Genes with Benjamini-Hochberg-adjusted p-values falling under the significance threshold of 0.05 were deemed as transcriptome-wide significant and were taken forward for further enrichment analyses. The union of genes significant to Salmon and STAR models formed the gene list entered gprofiler2 0.2 (24) gene ontology enrichment and the STRING 11.0 (25) protein-protein interaction network tools using default settings. The *p*-values returned by this enrichment analysis are adjusted for multiple testing by gprofiler2’s default gSCS algorithm. Additionally, a supplementary analysis was conducted with the Revised Amyotrophic Lateral Sclerosis Functional Rating Scale (ALSFRS-R) (26) lower limb subscore as the outcome, adjusting for age, sex and the estimated surrogate variables. This subscore combined scored answers for questions relating to the ability to walk and the ability to climb stairs, with lower values reflecting decreased ability.

Finally, the isoform-level length-scaled transcript counts generated by Salmon for the muscle data were analyzed through the IsoformSwitchAnalyzeR (27) 1.12 library in R. This package implements visualization and annotation functions on top of a DEXSeq, which currently implements DESeq2’s normalization and estimation approaches to iteratively compare the number of transcripts of a given exon compared to the rest of that gene (28) but in this case for isoforms. Isoforms were annotated to exons through the Gencode v32 primary assembly.

Following default prefiltering, isoforms were tested for differential usage using MND status as its condition, adjusted for age, sex and batch effect, then analyzed for alternative splicing as well as predicted the open reading frame (ORF) of each isoform of significantly switched genes. IsoFormSwitchAnalyzer implements multiple functions for the export of all isoform sequences for genes with a significant switch, and the import of the results from external annotation databases utilizing these. The databases were CPAT (using the suggested human coding threshold of 0.364) (29) for coding potential, SignalP (30) for predicting signal peptide sites, IUPred2A (31) to predict intrinsically disordered regions (IDRs) and Pfam (32) to predict protein domains.

## Results

Quality control of the RNAseq reads for each sample using PHRED scores indicated the quality was high, with the main batch containing mean scores at each base pair across reads and samples of > 30. However, the samples demonstrated some form of degradation (potentially from long-term storage), particularly for blood. Mean genome alignment over all blood and muscle samples through STAR was 54.4% and 87.6% respectively, while the proportion of those which were assigned to exons were 29.8% and 58.2% respectively. For Salmon, a mean of 74.1% of reads mapped to the transcriptome and were successfully assigned a feature in muscle, whereas this number was only 40% in blood. The proportions of the reads successfully assigned to a feature are plotted in **Figure S1** for STAR and **Figure S2** for Salmon.

A principal component analysis of the transcript levels revealed a differing pattern of variation between the blood and muscle analyses (**Figure 1**). The second principal component of both blood and muscle (explaining 7.2% and 8.4% of variance in gene expression, respectively), captured sex differences (correlation *r* = 0.90 with *p* = 4.9 × 10^−14^ for blood, and *r* = −0.89 with *p* = 2.6 × 10^−16^ for muscle). The first principal component in muscle (explaining 22.3% of expression variance) captured differences between cases and controls. The second principal component in muscle is significantly correlated with the lower limb subscore of the ALSFRS-R when estimating the PCA in case counts only (*r* = 0.46, *p* = 0.014). The relative log expression of the counts (centered and non-centered) passing QC is displayed within **Figure S3**. Similar observations with regards to the principal components and correlations apply to the counts as generated through STAR (**Figure S4**).

**Figure 1.**
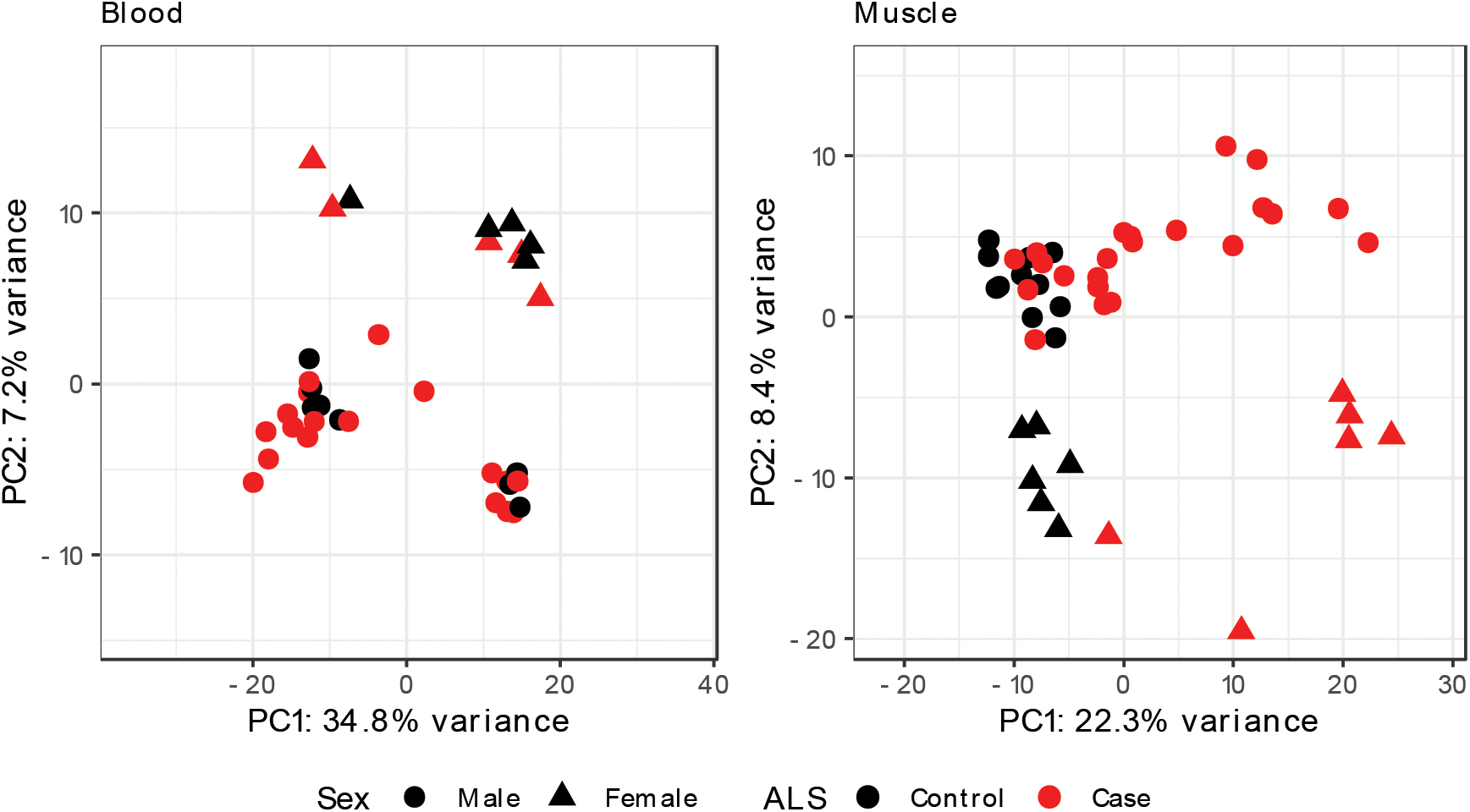
First two principal components of transcriptome log counts (regularized using Salmon) consisting of the top 500 most variable genes for blood and muscle respectively. PC1 in muscle captures variation between cases in sampled lower limb lower motor neurone scores (*r* = 0.40, *p* = 0.041), though does not fully separate cases and controls. PC2 captures sex differences for both tissue types (blood: *r* = 0.90, *p* = 4.9 × 10^−14^ ; muscle: *r* = −0.89, *p* = 2.6 × 10^−16^). Ellipses added to visualize confidence between cases and controls.

Testing for case-control differences in gene-expression levels in muscle while correcting for age, sex and replicate batch effect identified additional confounding within the data. The genomic inflation factor (GIF) of Salmon and STAR were 2.53 and 3.47 respectively, with 3258 and 6874 transcriptome-wide significant genes detected under the adjusted p-value of 0.05 (**Figure S5**). This was corrected with the addition of an automatically determined number of surrogate variables to each model as calculated by permutation procedure. With this correction the GIF falls to 1.18 and 1.13, with 38 and 26 significant genes for Salmon and STAR-aligned data respectively. For the blood transcriptome analysis, such inflation was not an issue (a GIF of 0.85 and 1.01 in Salmon and STAR respectively, with two significant genes each) and as such no SVs were added to the model.

Of the 38 and 26 genes detected by Salmon and STAR respectively, there was an intersection of 12 (**Table 1**) with the same direction of fold change for both methods. The complete list of transcriptome-wide genes for each model in each tissue type as well as their test statistics can be found in individual tabs of **Table S2**. Of the 52 genes identified by either Salmon and STAR models, 9 were long non-coding RNAs and 5 were pseudogenes. Of the 38 protein-coding genes, 7 had lower expression and 31 had higher expression in MND cases versus controls. As one of these genes, AHRR, is a known smoking biomarker in blood (33) we conducted an additional iteration of the ‘base’ model for muscle with the addition of ever-smoked status (23 never, 22 ever). MND status was found to be significantly associated with AHRR count in the STAR model (log_2_FC = 1.98, *p* = 0.0085), whereas smoking history was not (log_2_FC −0.11, *p* = 0.88).

**Table 1.**
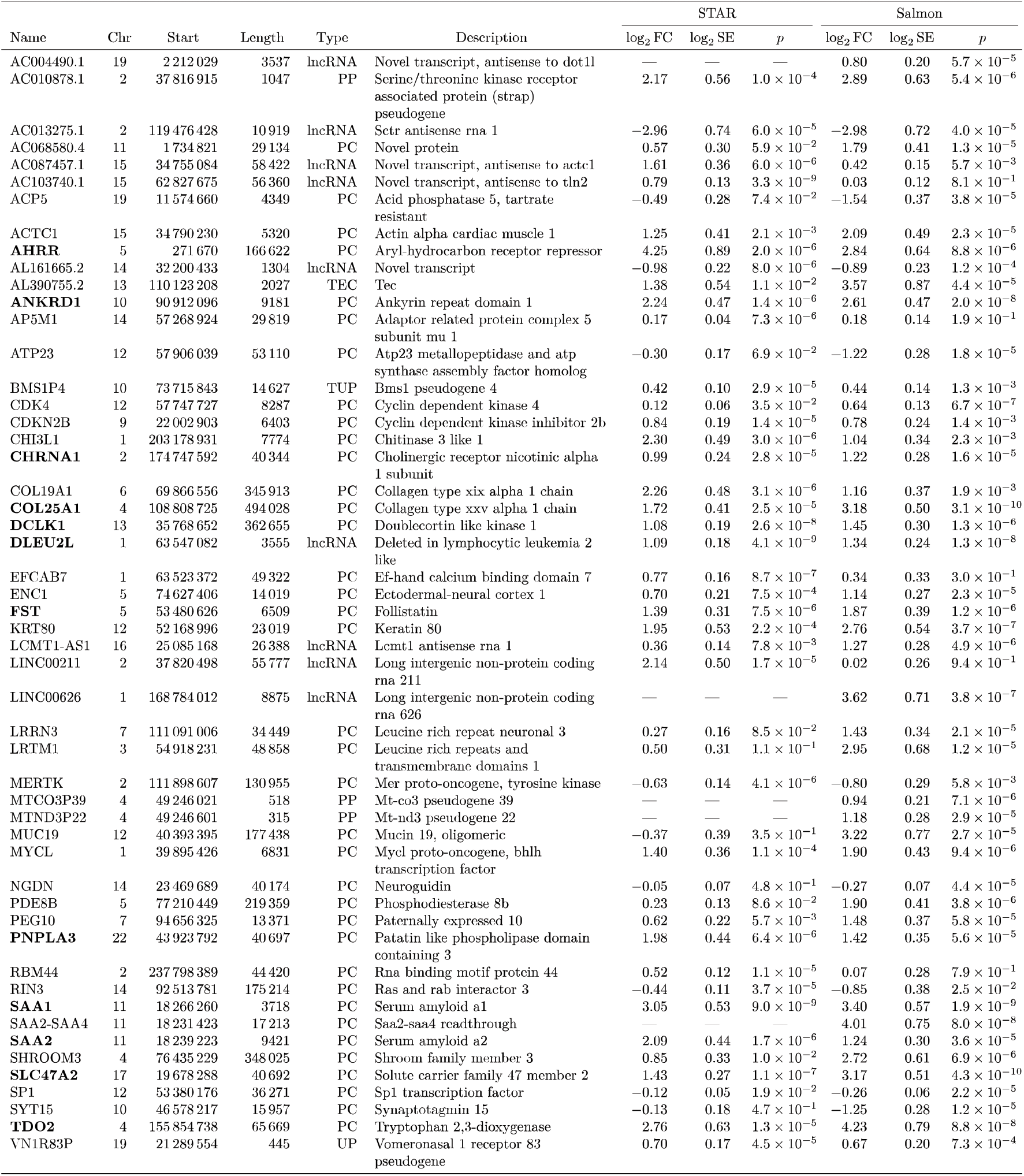
Overlap of transcriptome-wide significant differences between MND cases and controls from STAR and Salmon models with log2 fold change and Benjamini-Hochberg adjusted *p*-values. Bolded names are significant in both models. lncRNA: long non-coding RNA; PC: protein coding; PP: processed pseudogene; UP: Unprocessed pseudogene; TUP: transcribed unprocessed pseudogene.

A gene ontology enrichment analysis conducted on the results of each method (**Table 2**) revealed shared enrichment for the GO biological processes term of acute phase response (*p* = 1.4 × 10^−3^ and *p* = 5.5 × 10^−4^ for Salmon and STAR respectively), attributable to the components of serum amyloid A within the set. Other notable terms enriched in the significant results for Salmon were cardiac muscle tissue morphogenesis (*p* = 4.4 × 10^−3^), positive regulation of synapse assembly (*p* = 4.7 × 10^−3^), and tissue migration (*p* = 7.3 × 10^−3^). For STAR, notable terms were skeletal muscle tissue growth (*p* = 9.7 × 10^−3^), axonogenesis involved in innervation (*p* = 8.4 × 10^−3^) and neutrophil clearance (*p* = 7.0 × 10^−3^). However, these terms test as enriched due to having one or two significant genes detected from a small pool of other genes in the global set annotated with the term. The unified set of differentially expressed proteins between STAR and Salmon in muscle was identified to have significantly more interactions (9) than expected (4, *p* = 0.01) according to the STRING pathway analysis, indicating a likely biological relation if not interaction between them (**Figure S6**).

**Table 2.**
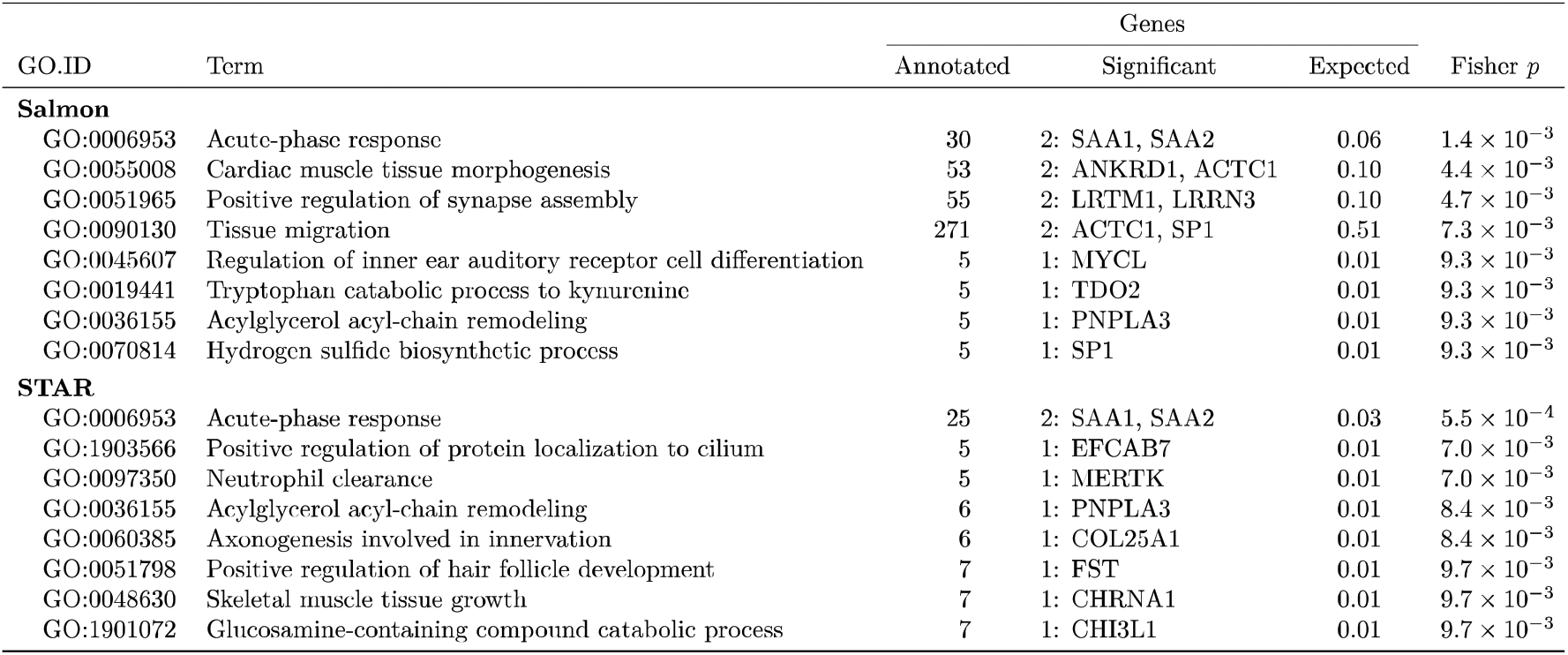
Enriched gene ontology (GO) terms for both Salmon and STAR models, as generated from TopGO. The total number of annotated genes from the greater set is followed by the number of those which are significantly differentially expressed, as well as the enrichment of the term as calculated by Fisher’s exact test. Significant/background genes are subset to those with GO terms (27/14475 for salmon and 19/13588 for STAR).

Within the blood sample, the *ODZ1* homolog pseudogene *AL0087071*, notably found on the X-chromosome, was the only gene differentially expressed by both Salmon and STAR methods (respective log_2_FCs of 3.14 and 3.07, with a *p* = 1.9 × 10^−6^ and *p* = 3.5 × 10^−6^, **Table S3**). *EDH2* was not detected by the STAR aligner though was significantly expressed in Salmon (log_2_FC of −1.75; *p* = 7.0 × 10^−7^) and the expression of *FSD1L* was only found significant in STAR (log_2_FC of 0.57; *p* = 2.0 × 10^−6^).

The results of the supplementary analyses on the 28 cases in predicting the ALSFRS-R lower limb subscores are reported in **Table S4**. The STAR model identified 11 genes significantly expressed in change of scoring and the Salmon model identified 10, with 3 genes (*IGFN1, PBX1*, and *TMEM9*) sharing association in both. Both models were adjusted for with two additional surrogate variables (as the number automatically suggested by the function).

A total of 17 genes were identified with isoform switches within the muscle sample falling underneath the FDR-adjusted p-value (q-value) of 0.05 and having predicted functional consequences to the expressed protein. The list of features, their direction of consequence and the genes falling under each is listed in **Table S5**. The individual per-isoform tests for each gene are available in **Table S6**. No significant (FDR < 0.05) directionality of functional consequences of these isoform differences was observed for any genomic feature, although there was a slight increase in switching to longer transcripts (8:6 longer vs. shorter switches) and an increase in exon number gain (6:3 gain vs. loss of exon count).

From the isoform switches with functional consequences, only *SAA2* was represented within our main gene-level analysis. *SAA2* featured an increase of its ENST00000256733 isoform fraction by 24% (*q* = 5.74 × 10-11) correlated with a decrease of its ENST00000528349 isoform fraction by 25% (*q* = 9.84 × 10^−11^). The resulting functional consequence was an overall decrease in transcript length by a shortened 3’ UTR and the reduction of intron 3-4 from 3090 to 394 bp, increasing its predicted coding potential (**Figure 2**). Two other isoform switches with predicted gain in coding potential are *PLCE1* (**Figure S7**) and *SAMD4A* (**Figure S8**). *PIP5K1B* (**Figure S9**) and *MYH3* (**Figure S10**) also have gain in domains which indicate that the isoform with increased expression encodes the more complete version of the protein.

**Figure 2.**
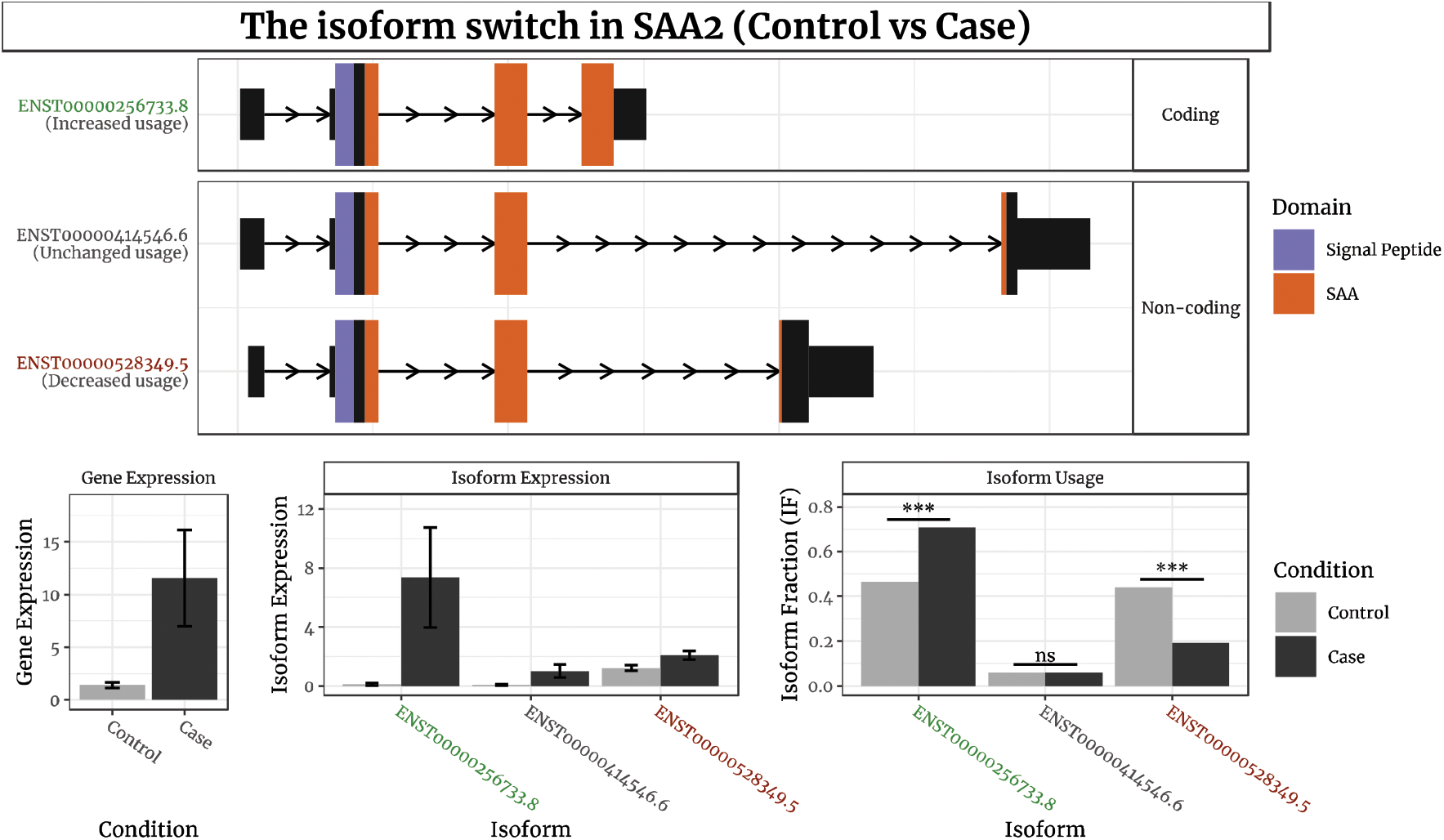
Visualization of isoform expression in the transcriptome-wide significant gene SAA2. The coloured plots at the top show three transcripts (detected via Salmon), annotated for their inclusion of protein domains as well as the site of their transcription start and termination sites. The top transcript is predicted to have high coding potential through CPAT. The bottom graphs individually delineate the difference in mean counts between cases and controls in the expression of the gene and its constituent isoforms. The rightmost graph indicates the isoform expression as a proportion of the total expression across all isoforms within cases and controls separately, indicating a greater proportion of expression for the coding variant and a decreased proportion of expression for the second non-coding variant.

## Discussion

The majority of studies on muscle changes associated with MND have been performed in mice (34,35) or iPSC-derived cell-culture (36). To our knowledge, our study is one of the first investigations into the transcriptomic machinery in human MND muscle tissue and provides a substantial increase in the number of individuals examined. We identify 38 protein-coding genes differentially expressed between MND cases and controls with 31 of these being upregulated in cases. Muscle biopsies were sampled from the healthier leg in MND cases.

Serum amyloid A transcripts (specifically *SAA1, SAA2* and the *SAA2-SAA4* readthrough, contiguously located on chromosome 11) were highly expressed in MND patients. SAA is persistently elevated in other chronic inflammatory diseases and a reliable indicator of inflammation due to an up-to 1000-fold increase in plasma levels during acute phase responses (37). In a heightened response, SAA most often becomes associated with the lipid surface of high-density lipoproteins (HDLs), possibly as a stabilized hub to interact with other cellular components (38). In its association with HDL cholesterol, SAA is indicated to be involved in stimulating the inflammatory response within the innate immune system (39). Other acute phase proteins such as C-reactive protein and CD14 have been detected as elevated in MND sera, and increased levels of these are reported to be associated with disease progression and mortality rate (40). In addition to the aforementioned acute phase proteins, several components of the complement system including C3 (41), C4 (42), and C5a/C5b-9 (43) have been reported to be higher in the whole blood, serum or plasma of ALS cases to controls in previous studies. As such, it may be of value in the future to investigate local accumulation of SAA as a biomarker for disease progression and as a representation of the overall inflammatory response.

Several genes associated with tissue generation are detected as positively expressed in cases. *FST* (follistatin) is a receptor agonist which binds to other transforming growth factor-β ligands, including myostatin and activin-A which would otherwise limit muscle mass without inhibition (44). Additionally, *FST* has an antagonistic effect on several bone morphogenic proteins, of which it forms a complex with and alters signaling to induce muscle hypertrophy while reducing atrophy (45). *MYH3*, a myosin heavy-chain encoding gene primarily expressed embryonically, was not captured by the gene-based analyses; however, the differential isoform expression analysis indicated a heightened expression of its most complete isoform within MND cases. This is of particular interest as *MYH3* has been indicated as a valid and robust biomarker for skeletal muscle tissue regeneration across all ages in other muscle-wasting diseases such as Duchenne muscular dystrophy (46). Muscle tissue does not exist in the body solely as actin filaments, but bundled within a complex net of connective tissue and infiltrating immune cells (47). Other genes instrumental in tissue development and differentiation found expressed higher in cases are the keratin *KRT80* (48), the actin-binding protein encoding *SHROOM3* which is utilized for epithelial folding in neural tube closure (49), and filamentous actin-binding ectodermal-neural cortex 1 *ENC1* (50).

Several transcripts found over-expressed in MND cases are not principally expressed by skeletal muscle tissues in adulthood. For instance, *ACTC1* was found to be significantly upregulated but not *ACTA1. ACTC1* encodes the α-cardiac muscle actin which constitutes < 5% of skeletal muscle by adulthood, whereas *ACTA1* encodes the almost-identical predominant muscular isoform α-skeletal muscle actin (51). Within a small case study of 7 congenital myopathy patients with a complete lack of α-skeletal muscle actin, α-cardiac actin was shown to constitute muscle tissues and the highest amount correlated to the participant with the best muscular function (52). However, as *ACTA1* has been previously identified to be increased as a function of compensatory hypertrophy in a small real-time PCR study of 5 ALS patients, a larger sample size may capture the increased expression of both it and *ACTC1* (53). *ANKRD1*/*CARP* is the ankryn repeat domain most present in cardiac tissue and is shown to be over-expressed within MND muscle compared to controls unlike its skeletal muscle paralog, *ANKRD2*/*ARPP*. However, *ANKRD1* expression has also been noted to increase in skeletal muscle with denervation-induced atrophy in murine models (54).

The expression of extracellular matrix components and collagens *COL19A1* and *COL25A1* was also found to be increased with muscle tissue genes such as *ACTC1. COL25A1*, encoding the collagen type XXV alpha 1 chain, is a known Alzheimer’s risk gene (55) for its role in assisting the formation of characteristic senile plaques when over-expressed in the brain (56). The cleavage of *COL25A1* creates the proteolytic CLAC (collagenous Alzheimer amyloid plaque component), and as such has the alias of *CLAC-P* (precursor) (57). Collagen XXV is expressed primarily in skeletal muscle during development, acting as a required factor in myofiber fusion from myoblasts in early myogenesis (58). *COL25A1*-KO mice nearly completely failed to innervate skeletal muscle tissue during development by not being able to elongate and branch motor axon bundles, indicating it as a required component to initial intramuscular motor innervation (59). Collagen XIX structures part of the basement membranes, and is present in a host of different tissues including skeletal muscle, axons, epithelia and brain intraneurons (60). As collagens are vital in the production of functional skeletal muscle tissue, their increased expression in MND further supports the evidence of attempted muscle repair as a compensatory response. Higher *COL19A1* expression levels in MND muscle biopsies have already been linked to faster progression of the disease (61).

A few other highly upregulated genes of note include some that are influenced by the multiple upregulated transcription factors in the set (*SP1* and *MYCL*). The increased expression of *TDO2* and *AHRR* may indicate the presence of prior *AHR* upregulation, which has been indicated to be invoked in neural cell proliferation (62). Following *AHRR* and the aforementioned *SAA1*, tryptophan catabolism enzyme *TDO2* has the largest positive fold-change in expression. *TDO2* is ordinarily expressed in the liver as a tryptophan regulator and is also known as an oncogene commonly expressed in most cancers (63). While the products of the catabolic kynurenine pathway include metabolites used for neurotransmission and immune regulation, *TDO2* depletes free tryptophan used for proliferation of T-cells and produces kynurenine that inhibits the proliferation of effector T lymphocytes and promoting regulatory T-cell differentiation, and activates the transcription factor *AHR* (64). The kynurenine pathway itself produces metabolites that influence energy homeostasis in skeletal muscle and other tissues, and kynurenic acid (a product of the pathway) increases energy utilization while also acting as an *AHR* ligand (65).

## Conclusions

In summary, we identify 52 genes whose expression in muscle is significantly associated with MND cases versus controls, with the majority showing increased expression in cases. These genes encode vital structural components of muscle, particularly in regenerative or early developmental processes. This provides novel insight into the repair mechanisms triggered in the presence of neurodegeneration and muscle wasting in MND, as well as the chronic inflammation within these tissues as evidenced by expression of acute-phase proteins.

## Supporting information

Primary Supplement

## Data Availability

The data that support the findings of this study are available on request from the corresponding author. The raw transcript data are not publicly available due to ethical restrictions; however, gene and transcript counts have been made available. The datasets generated during and/or analysed during the current study are available in the University of Queensland data collection repository.

https://doi.org/10.48610/b722f1f

## Abbreviations

*ALS-FRS-R*: Revised Amyotrophic Lateral Sclerosis Functional Rating Scale
*ALS*: Amyotrophic lateral sclerosis
*EM*: Expectation maximalization
*GIF*: Surrogate variable analysis
*lncRNA*: Long non-coding RNA
*MND*: Motor neurone disease
*NMJ*: Neuromuscular junction
*ORF*: Open reading frame
*PC*: Protein coding
*PCA*: Principal component analysis
*PP*: Processed pseudogene
*RIN*: RNA integrity number
*TUP*: Transcribed unprocessed pseudogene.
*UP*: Unprocessed pseudogene

## Declarations

### Ethics approval and content to participate

All work performed in this study was approved by the Royal Brisbane and Women’s Hospital and University of Queensland human research ethics committees. All participants provided written informed consent.

### Consent for publication

Not applicable.

### Availability of data and materials

The data that support the findings of this study are available on request from the corresponding author. The raw transcript data are not publicly available due to ethical restrictions; however, gene and transcript counts have been made available. The datasets generated during and/or analysed during the current study are available in the University of Queensland data collection repository, https://doi.org/10.48610/b722f1f.

## Authors’ contributions

A.F., A.F.M., and N.R.W. wrote the paper. A.F. conducted the analyses. A.F.M., N.R.W., and S.T.N. conceptualized this work. S.W and V.B. processed the samples for sequencing. L.M.W. and A.K.H. allocated laboratory resources and supervised sample processing. P.A.M. and R.D.H. provided samples. S.T.N. and F.J.S. performed the muscle biopsies. All authors read and approved the final manuscript.

## Acknowledgements

The authors thank all patients and participants who provided biological samples for use in this research.

## Supplemental Information

### Primary supplement

PDF. Figures S1-S10 and Table S1, S3-S5.

**Table S2**. Excel. Complete list of transcriptome-wide genes for each model in each tissue type as well as their test statistics.

**Table S6**. Excel. Differentially expressed isoforms as estimated from the IsoformSwitchAnalyzeR implementation of DEXSeq.

## References

1. Cappello V, Francolini M. Neuromuscular Junction Dismantling in Amyotrophic Lateral Sclerosis. IJMS. 2017 Oct 3;18(10):2092.

2. Manzano R, Toivonen JM, Moreno-Martínez L, Torre M, Moreno-García L, López-Royo T, et al. What skeletal muscle has to say in amyotrophic lateral sclerosis: Implications for therapy. Br J Pharmacol. 2021 Mar;178(6):1279–97.

3. Morris J. Amyotrophic Lateral Sclerosis (ALS) and Related Motor Neuron Diseases: An Overview. The Neurodiagnostic Journal. 2015 Sep;55(3):180–94.

4. Masrori P, Van Damme P. Amyotrophic lateral sclerosis: a clinical review. Eur J Neurol. 2020 Oct;27(10):1918–29.

5. Weddell T, Bashford J, Wickham A, Iniesta R, Chen M, Zhou P, et al. First-recruited motor units adopt a faster phenotype in amyotrophic lateral sclerosis. The Journal of Physiology [Internet]. 2021 Jul 14 [cited 2021 Jul 20];n/a(n/a). Available from: https://doi.org/10.1113/JP281310

6. Taylor JP, Brown RH, Cleveland DW. Decoding ALS: from genes to mechanism. Nature. 2016 Nov;539(7628):197–206.

7. Xue YC, Ng CS, Xiang P, Liu H, Zhang K, Mohamud Y, et al. Dysregulation of RNA-Binding Proteins in Amyotrophic Lateral Sclerosis. Front Mol Neurosci. 2020 May 29;13:78.

8. Tank EM, Figueroa-Romero C, Hinder LM, Bedi K, Archbold HC, Li X, et al. Abnormal RNA stability in amyotrophic lateral sclerosis. Nat Commun. 2018 Dec;9(1):2845.

9. Butti Z, Patten SA. RNA Dysregulation in Amyotrophic Lateral Sclerosis. Front Genet. 2019 Jan 22;9:712.

10. Dols-Icardo O, Montal V, Sirisi S, López-Pernas G, Cervera-Carles L, Querol-Vilaseca M, et al. Motor cortex transcriptome reveals microglial key events in amyotrophic lateral sclerosis. Neurol Neuroimmunol Neuroinflamm. 2020 Sep;7(5):e829.

11. van Rheenen W, Diekstra FP, Harschnitz O, Westeneng HJ, van Eijk KR, Saris CGJ, et al. Whole blood transcriptome analysis in amyotrophic lateral sclerosis: A biomarker study. Raoul C, editor. PLoS ONE. 2018 Jun 25;13(6):e0198874.

12. Nijssen J, Aguila J, Hoogstraaten R, Kee N, Hedlund E. Axon-Seq Decodes the Motor Axon Transcriptome and Its Modulation in Response to ALS. Stem Cell Reports. 2018 Dec;11(6):1565–78.

13. Barham C, Fil D, Byrum SD, Rahmatallah Y, Glazko G, Kiaei M. RNA-Seq Analysis of Spinal Cord Tissues from hPFN1G118V Transgenic Mouse Model of ALS at Pre-symptomatic and End-Stages of Disease. Scientific Reports. 2018 Sep 13;8(1):13737.

14. Steyn FJ, Li R, Kirk SE, Tefera TW, Xie TY, Tracey TJ, et al. Altered skeletal muscle glucose- fatty acid flux in amyotrophic lateral sclerosis. Brain Communications. 2020 Sep 24;fcaa154.

15. Brooks BR, Miller RG, Swash M, Munsat TL. El Escorial revisited: Revised criteria for the diagnosis of amyotrophic lateral sclerosis. null. 2000 Jan 1;1(5):293–9.

16. Chen S, Zhou Y, Chen Y, Gu J. fastp: an ultra-fast all-in-one FASTQ preprocessor. Bioinformatics. 2018 Sep 1;34(17):i884–90.

17. Dobin A, Davis CA, Schlesinger F, Drenkow J, Zaleski C, Jha S, et al. STAR: ultrafast universal RNA-seq aligner. Bioinformatics. 2013 Jan;29(1):15–21.

18. Liao Y, Smyth GK, Shi W. The R package Rsubread is easier, faster, cheaper and better for alignment and quantification of RNA sequencing reads. Nucleic Acids Research. 2019 May 7;47(8):e47–e47.

19. Patro R, Duggal G, Love MI, Irizarry RA, Kingsford C. Salmon provides fast and bias-aware quantification of transcript expression. Nat Methods. 2017 Apr;14(4):417–9.

20. Soneson C, Love MI, Robinson MD. Differential analyses for RNA-seq: transcript-level estimates improve gene-level inferences. F1000Res. 2015;4:1521.

21. Anders S, Huber W. Differential expression of RNA-Seq data at the gene level – the DESeq package. :24.

22. Devlin B, Roeder K. Genomic Control for Association Studies. Biometrics. 1999;55(4):997–1004.

23. Leek JT, Johnson WE, Parker HS, Jaffe AE, Storey JD. The sva package for removing batch effects and other unwanted variation in high-throughput experiments. Bioinformatics. 2012 Mar 15;28(6):882–3.

24. Raudvere U, Kolberg L, Kuzmin I, Arak T, Adler P, Peterson H, et al. g:Profiler: a web server for functional enrichment analysis and conversions of gene lists (2019 update). Nucleic Acids Research. 2019 Jul 2;47(W1):W191–8.

25. Szklarczyk D, Gable AL, Lyon D, Junge A, Wyder S, Huerta-Cepas J, et al. STRING v11: protein–protein association networks with increased coverage, supporting functional discovery in genome-wide experimental datasets. Nucleic Acids Research. 2019 Jan 8;47(D1):D607–13.

26. Cedarbaum JM, Stambler N, Malta E, Fuller C, Hilt D, Thurmond B, et al. The ALSFRS-R: a revised ALS functional rating scale that incorporates assessments of respiratory function. Journal of the Neurological Sciences. 1999 Oct 31;169(1):13–21.

27. Vitting-Seerup K, Sandelin A. IsoformSwitchAnalyzeR: analysis of changes in genome- wide patterns of alternative splicing and its functional consequences. Bioinformatics. 2019 Nov 1;35(21):4469–71.

28. Anders S, Reyes A, Huber W. Detecting differential usage of exons from RNA-seq data. Genome Research. 2012 Oct 1;22(10):2008–17.

29. Wang L, Park HJ, Dasari S, Wang S, Kocher JP, Li W. CPAT: Coding-Potential Assessment Tool using an alignment-free logistic regression model. Nucleic Acids Research. 2013 Apr 1;41(6):e74–e74.

30. Almagro Armenteros JJ, Tsirigos KD, Sønderby CK, Petersen TN, Winther O, Brunak S, et al. SignalP 5.0 improves signal peptide predictions using deep neural networks. Nature Biotechnology. 2019 Apr 1;37(4):420–3.

31. Mészáros B, Erdős G, Dosztányi Z. IUPred2A: context-dependent prediction of protein disorder as a function of redox state and protein binding. Nucleic Acids Research. 2018 Jul 2;46(W1):W329–37.

32. Mistry J, Chuguransky S, Williams L, Qureshi M, Salazar GA, Sonnhammer ELL, et al. Pfam: The protein families database in 2021. Nucleic Acids Research. 2021 Jan 8;49(D1):D412–9.

33. Reynolds LM, Wan M, Ding J, Taylor JR, Lohman K, Su D, et al. DNA Methylation of the Aryl Hydrocarbon Receptor Repressor Associations With Cigarette Smoking and Subclinical Atherosclerosis. Circulation: Cardiovascular Genetics. 2015 Oct 1;8(5):707–16.

34. Ehmsen JT, Kawaguchi R, Mi R, Coppola G, Höke A. Longitudinal RNA-Seq analysis of acute and chronic neurogenic skeletal muscle atrophy. Scientific Data. 2019 Sep 24;6(1):179.

35. Gonzalez de Aguilar JL, Niederhauser-Wiederkehr C, Halter B, De Tapia M, Di Scala F, Demougin P, et al. Gene profiling of skeletal muscle in an amyotrophic lateral sclerosis mouse model. Physiological Genomics. 2008 Jan 1;32(2):207–18.

36. Lynch EM, Robertson S, FitzGibbons C, Reilly M, Switalski C, Eckardt A, et al. Transcriptome analysis using patient iPSC-derived skeletal myocytes: Bet1L as a new molecule possibly linked to neuromuscular junction degeneration in ALS. Experimental Neurology. 2021 Nov 1;345:113815.

37. Ye RD, Sun L. Emerging functions of serum amyloid A in inflammation. Journal of Leukocyte Biology. 2015 Dec;98(6):923–9.

38. Webb NR. High-Density Lipoproteins and Serum Amyloid A (SAA). Curr Atheroscler Rep. 2021 Feb;23(2):7.

39. Sack GH. Serum amyloid A – a review. Mol Med. 2018 Dec;24(1):46.

40. Beers DR, Zhao W, Neal DW, Thonhoff JR, Thome AD, Faridar A, et al. Elevated acute phase proteins reflect peripheral inflammation and disease severity in patients with amyotrophic lateral sclerosis. Sci Rep. 2020 Dec;10(1):15295.

41. Chen X, Feng W, Huang R, Guo X, Chen Y, Zheng Z, et al. Evidence for peripheral immune activation in amyotrophic lateral sclerosis. Journal of the Neurological Sciences. 2014 Dec 15;347(1):90–5.

42. Kjældgaard AL, Pilely K, Olsen KS, Øberg Lauritsen A, Wørlich Pedersen S, Svenstrup K, et al. Complement Profiles in Patients with Amyotrophic Lateral Sclerosis: A Prospective Observational Cohort Study. J Inflamm Res. 2021;14:1043–53.

43. Mantovani S, Gordon R, Macmaw JK, Pfluger CMM, Henderson RD, Noakes PG, et al. Elevation of the terminal complement activation products C5a and C5b-9 in ALS patient blood. J Neuroimmunol. 2014 Nov 15;276(1–2):213–8.

44. Lee SJ, Lee YS, Zimmers TA, Soleimani A, Matzuk MM, Tsuchida K, et al. Regulation of muscle mass by follistatin and activins. Mol Endocrinol. 2010/09/01 ed. 2010 Oct;24(10):1998–2008.

45. Winbanks CE, Chen JL, Qian H, Liu Y, Bernardo BC, Beyer C, et al. The bone morphogenetic protein axis is a positive regulator of skeletal muscle mass. Journal of Cell Biology. 2013 Oct 21;203(2):345–57.

46. Guiraud S, Edwards B, Squire SE, Moir L, Berg A, Babbs A, et al. Embryonic myosin is a regeneration marker to monitor utrophin-based therapies for DMD. Human Molecular Genetics [Internet]. 2018 Oct 9 [cited 2022 Feb 24]; Available from: https://academic.oup.com/hmg/advance-article/doi/10.1093/hmg/ddy353/5124574

47. Mukund K, Subramaniam S. Skeletal muscle: A review of molecular structure and function, in health and disease. WIREs Systems Biology and Medicine. 2020 Jan 1;12(1):e1462.

48. Li C, Liu X, Liu Y, Liu X, Wang R, Liao J, et al. Keratin 80 promotes migration and invasion of colorectal carcinoma by interacting with PRKDC via activating the AKT pathway. Cell Death Dis. 2018 Oct;9(10):1009.

49. Haigo SL, Hildebrand JD, Harland RM, Wallingford JB. Shroom Induces Apical Constriction and Is Required for Hingepoint Formation during Neural Tube Closure. Current Biology. 2003 Dec;13(24):2125–37.

50. Cui Y, Yang J, Bai Y, Li Q, Yao Y, Liu C, et al. ENC1 Facilitates Colorectal Carcinoma Tumorigenesis and Metastasis via JAK2/STAT5/AKT Axis-Mediated Epithelial Mesenchymal Transition and Stemness. Front Cell Dev Biol. 2021 Mar 16;9:616887.

51. Sztal TE, McKaige EA, Williams C, Ruparelia AA, Bryson-Richardson RJ. Genetic compensation triggered by actin mutation prevents the muscle damage caused by loss of actin protein. Stainier DYR, editor. PLoS Genet. 2018 Feb 8;14(2):e1007212.

52. Nowak KJ, Sewry CA, Navarro C, Squier W, Reina C, Ricoy JR, et al. Nemaline myopathy caused by absence of α-skeletal muscle actin. Ann Neurol. 2007 Feb;61(2):175–84.

53. Jensen L, Jørgensen LH, Bech RD, Frandsen U, Schrøder HD. Skeletal Muscle Remodelling as a Function of Disease Progression in Amyotrophic Lateral Sclerosis. Biomed Res Int. 2016;2016:5930621.

54. Laure L, Suel L, Roudaut C, Bourg N, Ouali A, Bartoli M, et al. Cardiac ankyrin repeat protein is a marker of skeletal muscle pathological remodelling: Cardiac ankyrin repeat protein in muscle plasticity. FEBS Journal. 2009 Feb;276(3):669–84.

55. Forsell C, Björk BF, Lilius L, Axelman K, Fabre SF, Fratiglioni L, et al. Genetic association to the amyloid plaque associated protein gene COL25A1 in Alzheimer’s disease. Neurobiol Aging. 2010 Mar;31(3):409–15.

56. Tong Y, Xu Y, Scearce-Levie K, Ptáček LJ, Fu YH. COL25A1 triggers and promotes Alzheimer’s disease-like pathology in vivo. Neurogenetics. 2010 Feb;11(1):41–52.

57. Hashimoto T, Fujii D, Naka Y, Kashiwagi-Hakozaki M, Matsuo Y, Matsuura Y, et al. Collagenous Alzheimer amyloid plaque component impacts on the compaction of amyloid-β plaques. acta neuropathol commun. 2020 Dec;8(1):212.

58. Gonçalves TJM, Boutillon F, Lefebvre S, Goffin V, Iwatsubo T, Wakabayashi T, et al. Collagen XXV promotes myoblast fusion during myogenic differentiation and muscle formation. Sci Rep. 2019 Dec;9(1):5878.

59. Tanaka T, Wakabayashi T, Oizumi H, Nishio S, Sato T, Harada A, et al. CLAC-P/Collagen Type XXV Is Required for the Intramuscular Innervation of Motoneurons during Neuromuscular Development. Journal of Neuroscience. 2014 Jan 22;34(4):1370–9.

60. Calvo AC, Moreno L, Moreno L, Toivonen JM, Manzano R, Molina N, et al. Type XIX collagen: a promising biomarker from the basement membranes. Neural Regen Res. 2020 Jun;15(6):988–95.

61. Calvo AC, Cibreiro GA, Merino PT, Roy JF, Galiana A, Rufián AJ, et al. Collagen XIX Alpha 1 Improves Prognosis in Amyotrophic Lateral Sclerosis. Aging and disease. 2019;10(2):278.

62. Juricek L, Coumoul X. The Aryl Hydrocarbon Receptor and the Nervous System. IJMS. 2018 Aug 24;19(9):2504.

63. Hoffmann D, Dvorakova T, Stroobant V, Bouzin C, Daumerie A, Solvay M, et al. Tryptophan 2,3-Dioxygenase Expression Identified in Human Hepatocellular Carcinoma Cells and in Intratumoral Pericytes of Most Cancers. Cancer Immunol Res. 2020 Jan;8(1):19–31.

64. Opitz CA, Somarribas Patterson LF, Mohapatra SR, Dewi DL, Sadik A, Platten M, et al. The therapeutic potential of targeting tryptophan catabolism in cancer. Br J Cancer. 2020 Jan 7;122(1):30–44.

65. Liu JJ, Movassat J, Portha B. Emerging role for kynurenines in metabolic pathologies. Current Opinion in Clinical Nutrition & Metabolic Care [Internet]. 2019;22(1). Available from: https://journals.lww.com/co-clinicalnutrition/Fulltext/2019/01000/Emerging_role_for_kynurenines_in_metabolic.15.aspx

