## Supplementary material for "RNA-seq analysis of skeletal muscle in motor neurone disease cases and controls": Primary Supplement

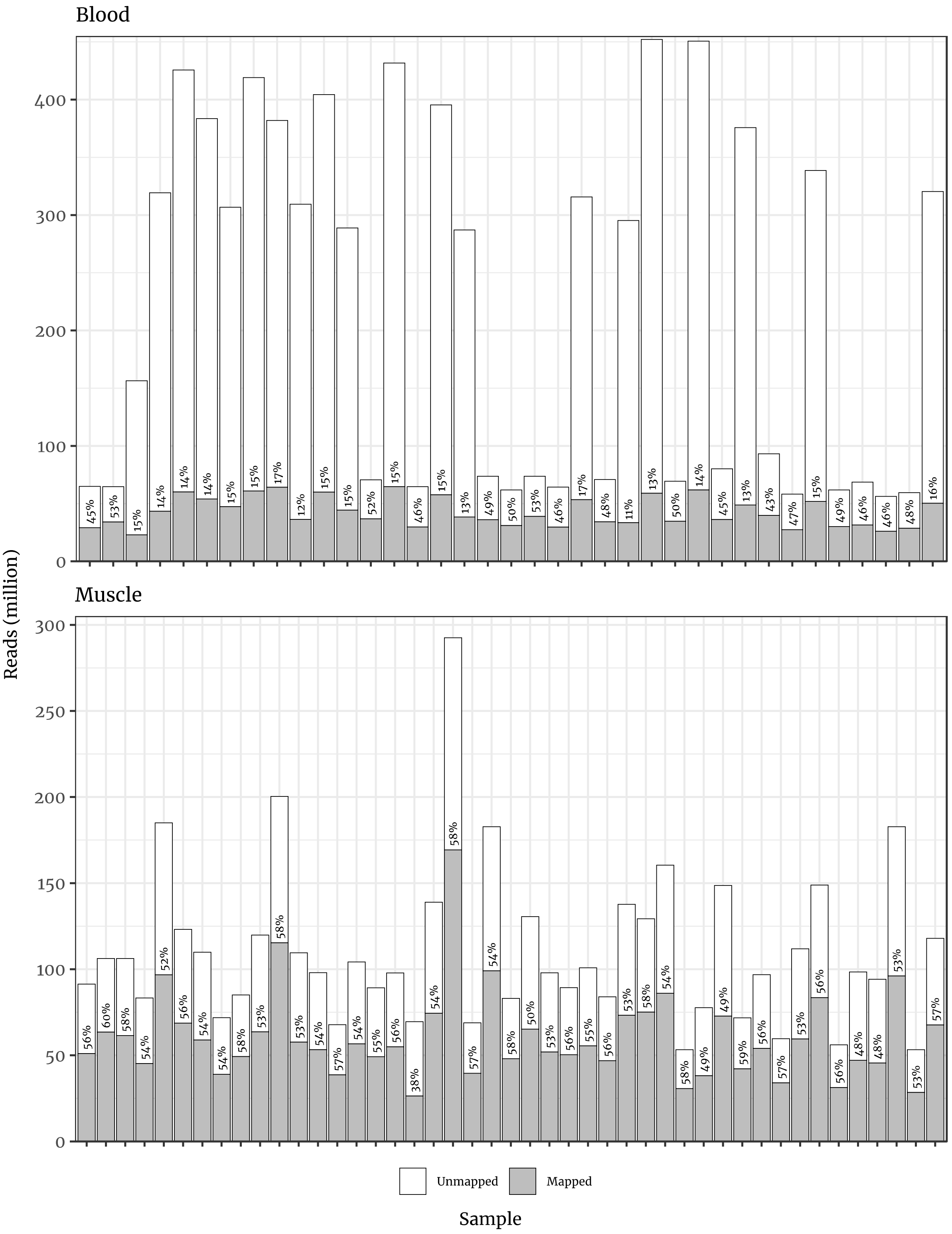


**Figure S1**. Per-individual quantification of read alignment to the genome using the STAR algorithm, across blood and muscle samples. The total number of reads counted for each sample is accompanied with a shaded proportion and a written percentage of reads that successfully and uniquely mapped to the transcriptome.

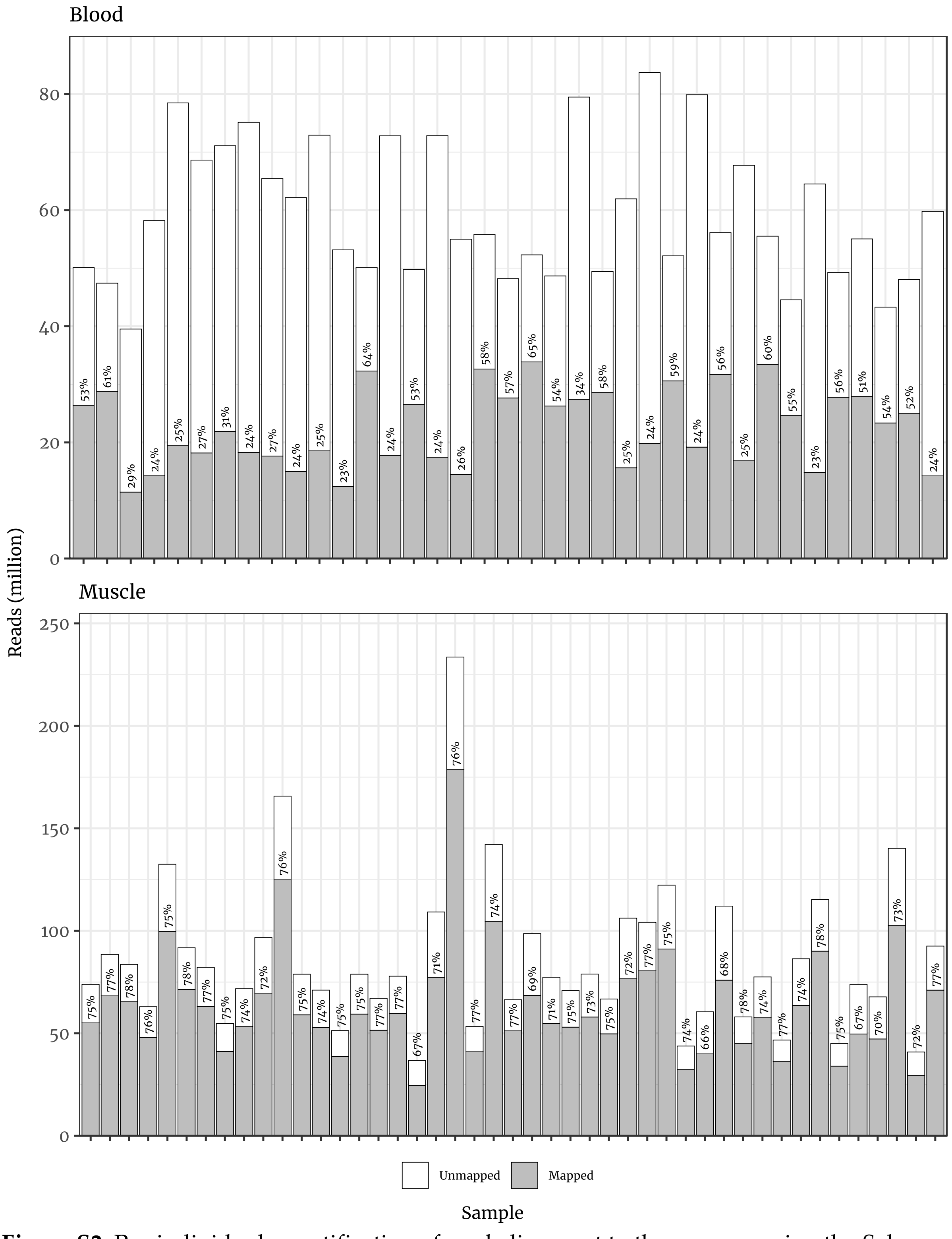


**Figure S2**. Per-individual quantification of read alignment to the genome using the Salmon algorithm, across blood and muscle samples. The total number of reads counted for each sample is accompanied with a shaded proportion and a written percentage of reads that successfully and uniquely mapped to the transcriptome.

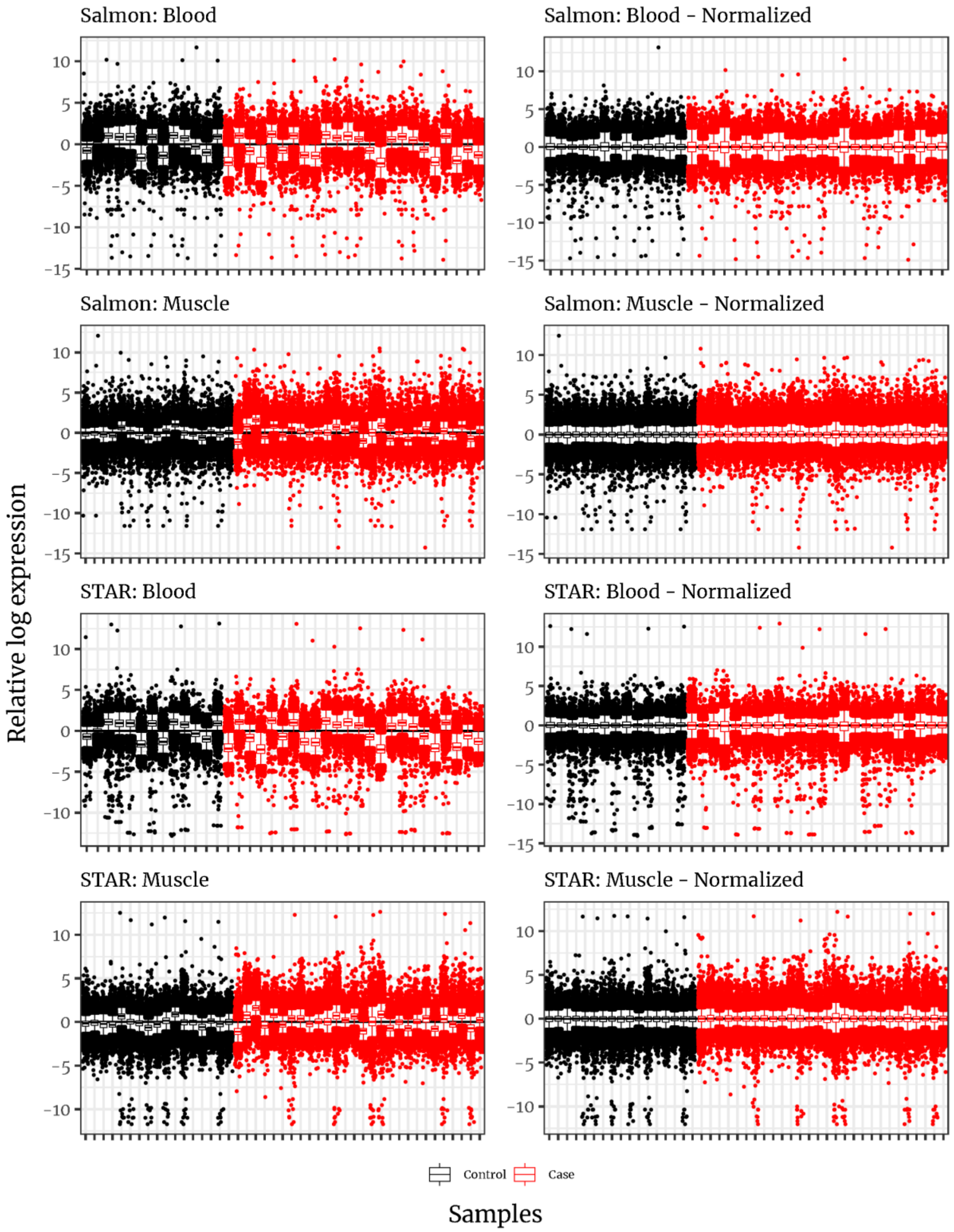


**Figure S3**. Relative log expression (RLE) of transcripts generated by Salmon mapping and STAR alignment to transcriptome and genome respectively, across samples. Normalized counts are generated using DESeq2-based centring.

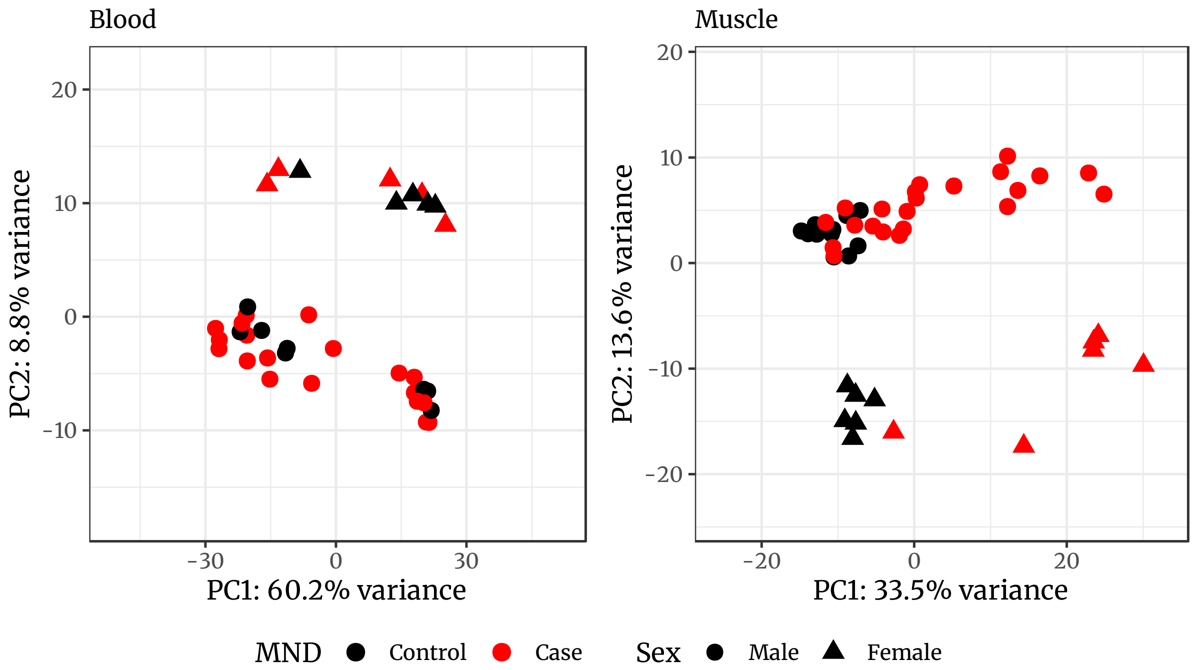


**Figure S4**. First two principal components of STAR transcriptome regularized log counts consisting of the top 500 most variable genes for blood and muscle respectively. PC1 in muscle captures variation between cases in lower limb lower motor neurone scores ($r = 0.42$, $p = 0.035$), though does not fully separate cases and controls. PC2 captures sex differences for both tissue types (blood: $r = 0.93$, $p = 1.0 \times 10^{-16}$; muscle: $r = -0.94$, $p = 2.6 \times 10^{-21}$). Ellipses added to visualize confidence between cases and controls.

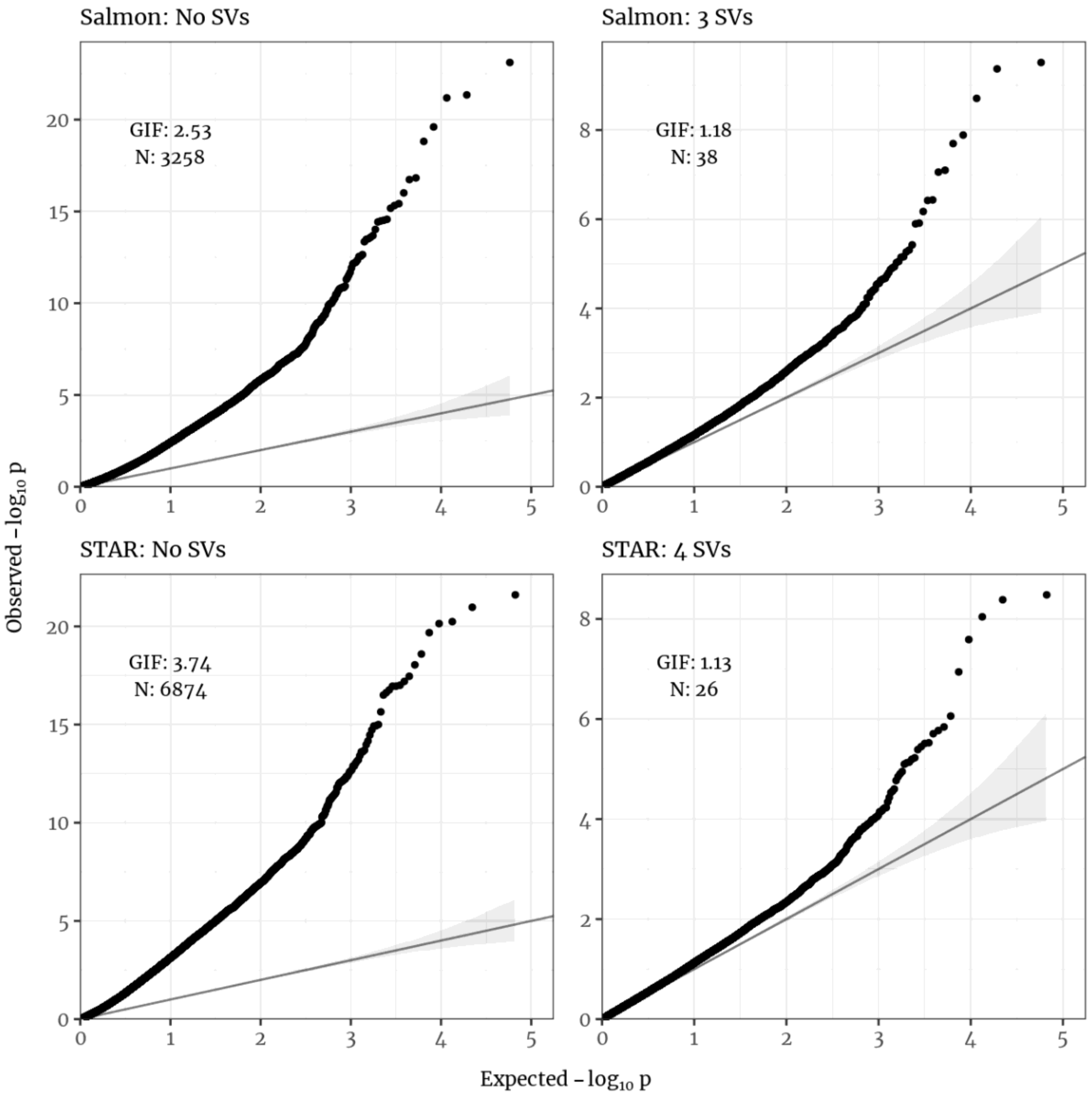


**Figure S5**. QQ plots of the DESeq2 muscle models as quantified from Salmon and STAR counts, annotated for number of transcripts passing the transcriptome-wide significant level and the model's genomic inflation factor (GIF). The "base" models are reported alongside the model with additional surrogate variables, the number of which was automatically selected while preserving MND status.

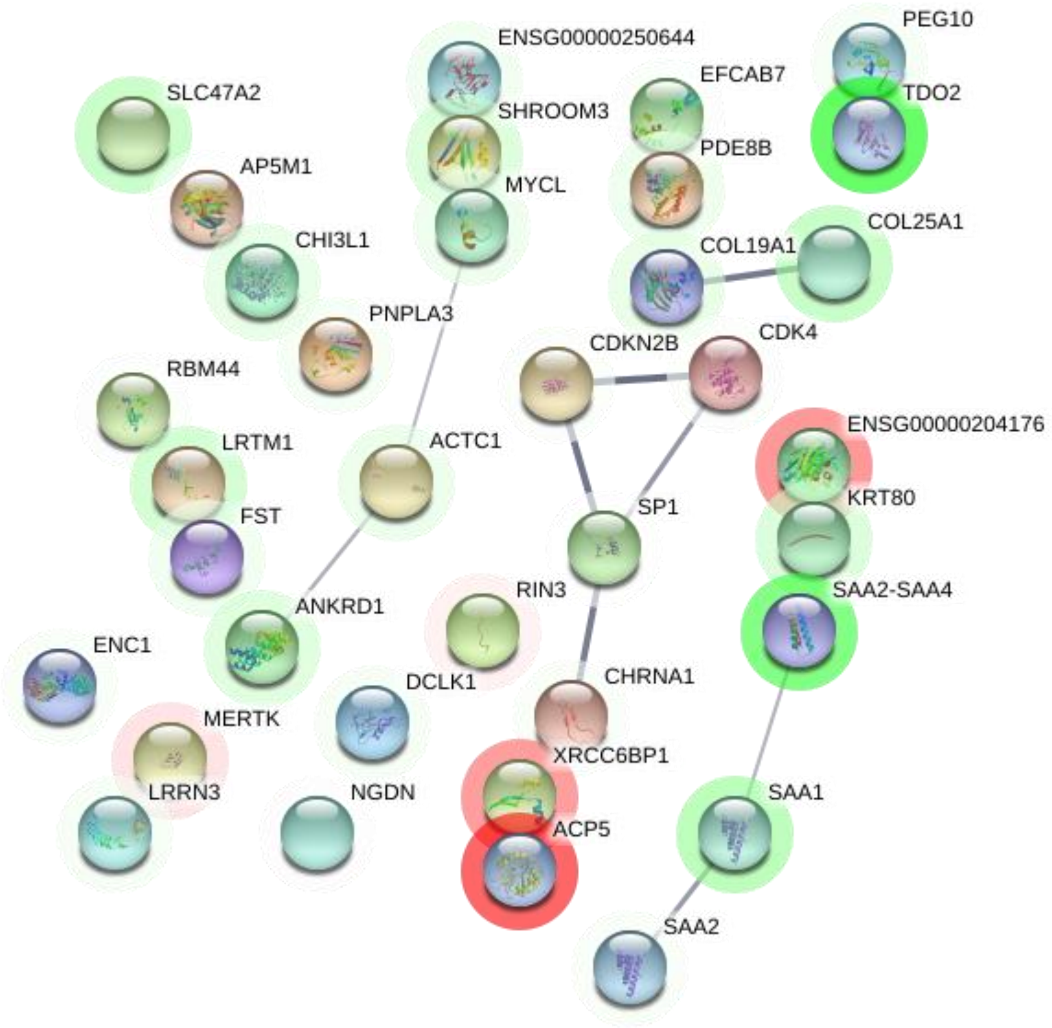


**Figure S6**. STRING-supported interactions between all significant protein-coding genes found in either Salmon and STAR models for muscle (35 in database). Thickness of lines indicates confidence in the interaction. Coloured halos indicate the direction and magnitude of log2 fold change from red (lower expression in MND) to green (higher expression in MND). Genes represented in both the Salmon and STAR results use their Salmon log2 fold change.

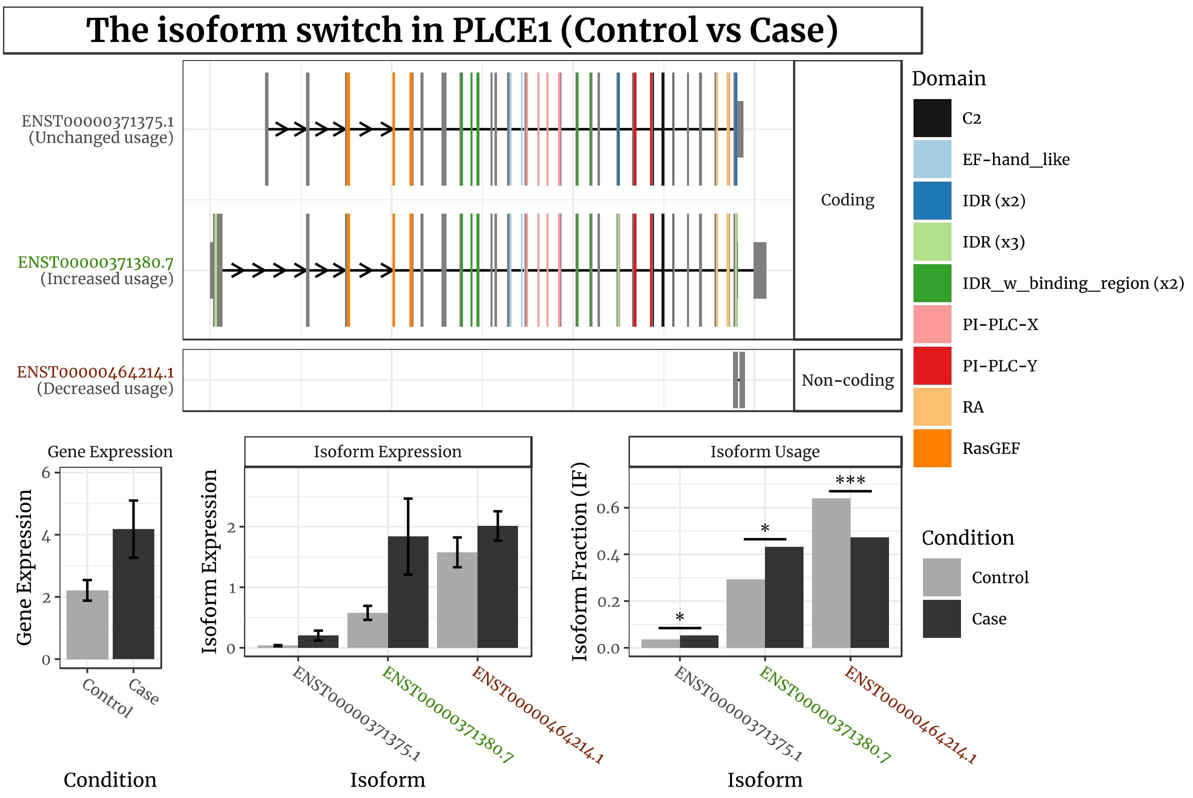


**Figure S7**. Visualization of isoform expression in the transcriptome-wide significant gene PLCE1. The coloured plots at the top show transcripts (detected via Salmon), annotated for their inclusion of protein domains as well as the site of their transcription start and termination sites. The top two transcripts are predicted to have high coding potential through CPAT. The bottom graphs individually delineate the difference in mean counts between cases and controls in the expression of the gene and its constituent isoforms. The rightmost graph indicates the isoform expression as a proportion of the total expression across all isoforms within cases and controls separately, indicating a greater proportion of expression for the longest coding variant and a decreased proportion of expression for the non-coding variant.

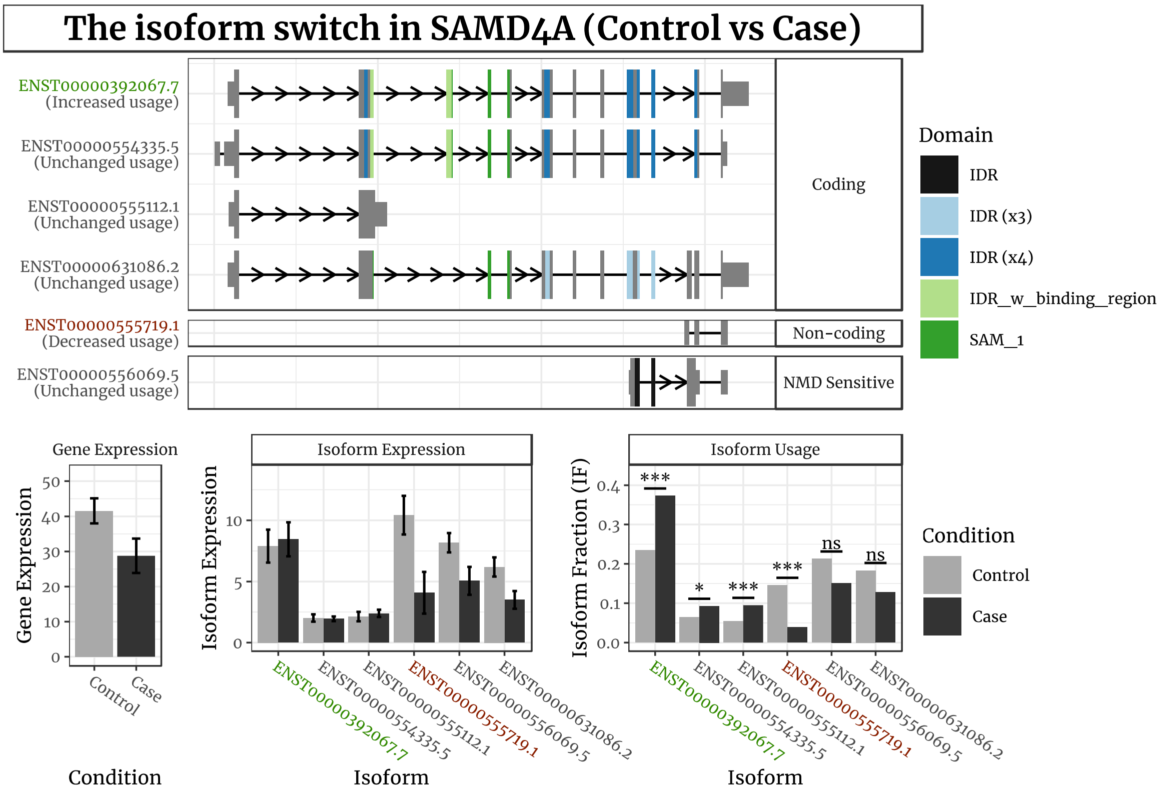


**Figure S8**. Visualization of isoform expression in the transcriptome-wide significant gene SAMD4A. The coloured plots at the top show transcripts (detected via Salmon), annotated for their inclusion of protein domains as well as the site of their transcription start and termination sites. The top four transcripts are predicted to have high coding potential through CPAT. The bottom graphs individually delineate the difference in mean counts between cases and controls in the expression of the gene and its constituent isoforms. The rightmost graph indicates the isoform expression as a proportion of the total expression across all isoforms within cases and controls separately, indicating a greater proportion of expression for the coding variant and a decreased proportion of expression for the non-coding variant.

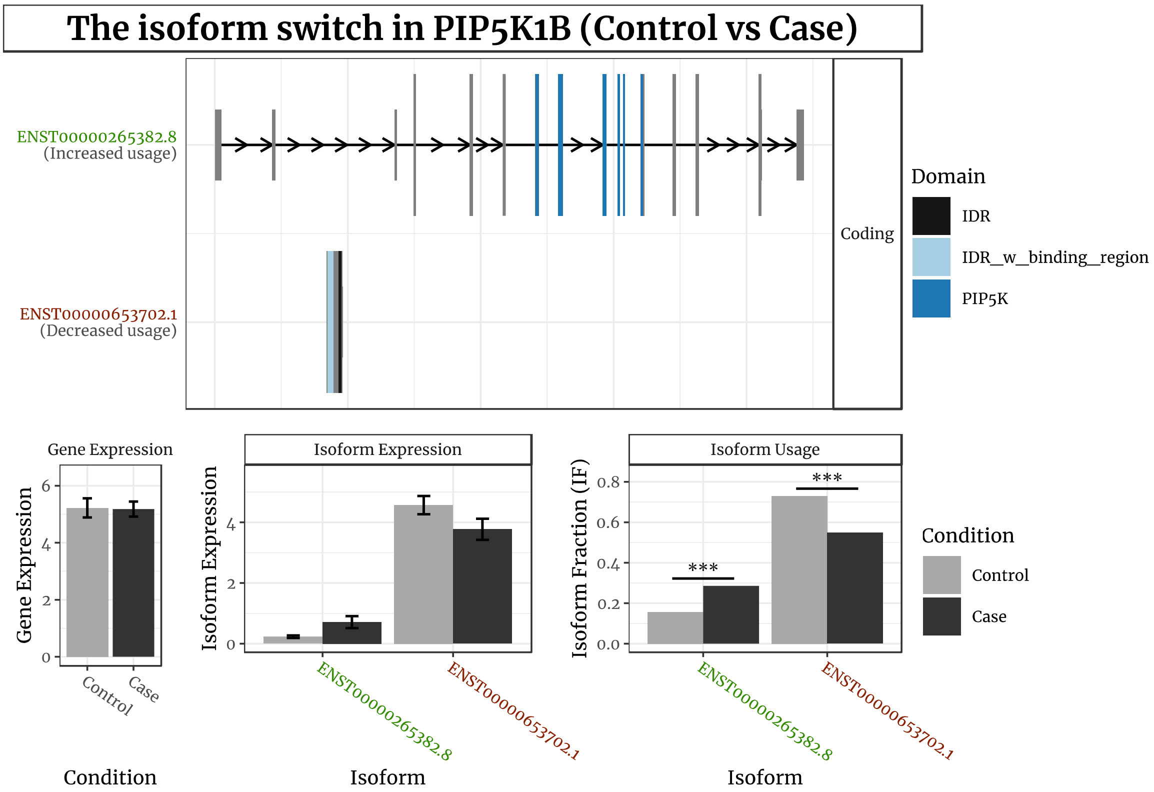


**Figure S9**. Visualization of isoform expression in the transcriptome-wide significant gene PIP5K1B. The coloured plots at the top show two transcripts (detected via Salmon), annotated for their inclusion of protein domains as well as the site of their transcription start and termination sites. The top transcript is predicted to have high coding potential through CPAT. The bottom graphs individually delineate the difference in mean counts between cases and controls in the expression of the gene and its constituent isoforms. The rightmost graph indicates the isoform expression as a proportion of the total expression across all isoforms within cases and controls separately, indicating a greater proportion of expression for the coding variant and a decreased proportion of expression for the non-coding variant.

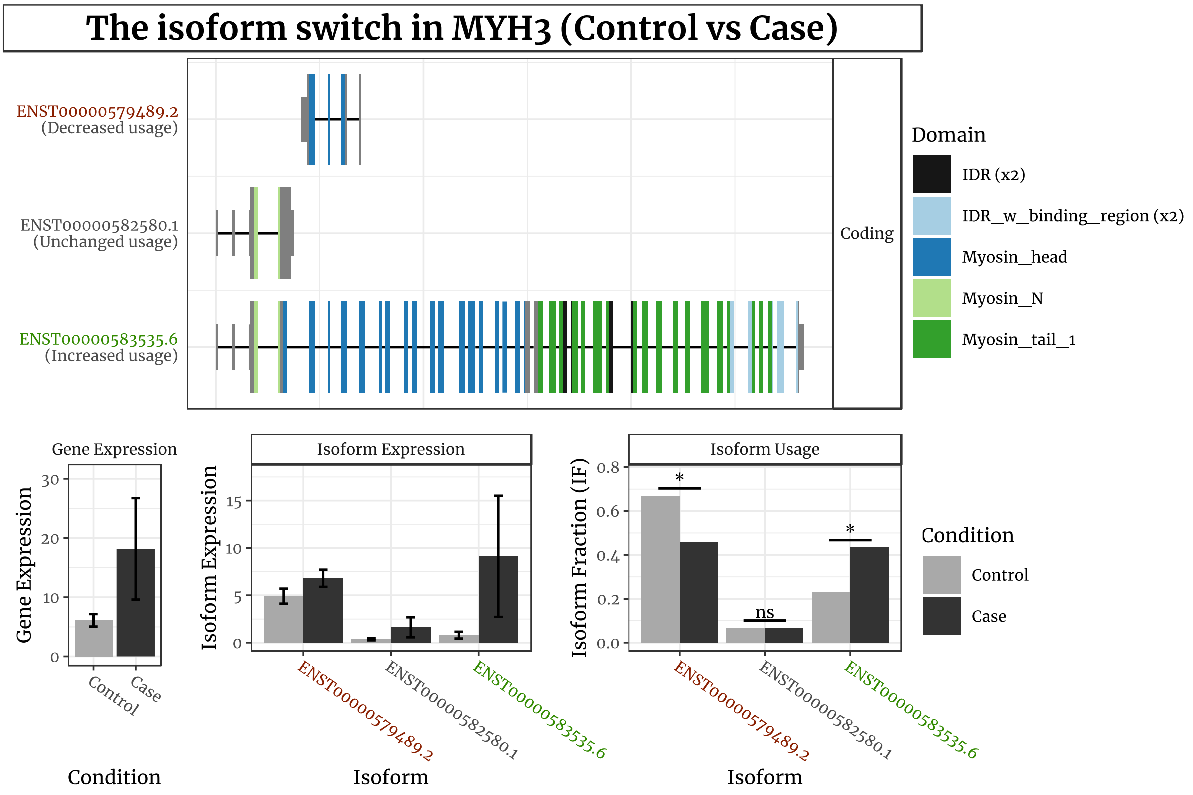


**Figure S10**. Visualization of isoform expression in the transcriptome-wide significant gene MYH3. The coloured plots at the top show three transcripts (detected via Salmon), annotated for their inclusion of protein domains as well as the site of their transcription start and termination sites. The three transcripts shown are predicted to have high coding potential through CPAT. The bottom graphs individually delineate the difference in mean counts between cases and controls in the expression of the gene and its constituent isoforms. The rightmost graph indicates the isoform expression as a proportion of the total expression across all isoforms within cases and controls separately, indicating a greater proportion of expression for the longest coding variant and a decreased proportion of expression for the shorter variants.

**Table S1**. Counts and description of the samples passing QC for muscle and blood, additionally stratified by MND status with totals. The $p$-value for age was estimated from ANOVA, whereas the other categorical variables were estimated from a chi-squared test.

| | Muscle | | | Blood | | |
|---|---|---|---|---|---|---|
| | Controls | Cases | $p$ | Controls | Cases | $p$ |
| $n$ | 17 | 28 | | 13 | 24 | |
| Age (mean (SD)) | 56.2 (15.0) | 59.4 (8.9) | 0.37 | 54.8 (16.5) | 60.2 (9.1) | 0.21 |
| **Sex** ($n$ (%)) | | | 0.50 | | | 0.44 |
| Male | 11 (64.7) | 22 (78.6) | | 8 (61.5) | 19 (79.2) | |
| Female | 6 (35.3) | 6 (21.4) | | 5 (38.5) | 5 (20.8) | |
| **Smoking** ($n$ (%)) | | | 0.13 | | | 0.29 |
| Ex- | 4 (23.5) | 14 (50.0) | | 3 (25.0) | 10 (41.7) | |
| Never | 12 (70.6) | 11 (39.3) | | 9 (75.0) | 12 (50.0) | |
| Current | 1 (5.9) | 3 (10.7) | | 0 (0.0) | 2 (8.3) | |
| **Batch** ($n$ (%)) | | | 0.90 | | | — |
| First | 2 (11.8) | 5 (17.9) | | | | |
| Second | 15 (88.2) | 23 (82.1) | | | | |

**Table S3**. Transcriptome-wide significant genes for blood with log2 fold change and Benjamini-Hochberg adjusted p-values reported for STAR and Salmon. PC: protein coding; PP: processed pseudogene.

| | | | | | | STAR | | salmon | |
|---|---|---|---|---|---|---|---|---|---|
| Name | Chr | Start | Length | Type | Description | $\log_2$ FC | $p$ | $\log_2$ FC | $p$ |
| AL008707.1 | X | 125 203 805 | 534 | PP | Odz, odd Oz/ten-m homolog 1 (Drosophila) (ODZ1) pseudogene | 3.14 | $1.9 \times 10^{-6}$ | 3.07 | $3.5 \times 10^{-6}$ |
| EHD2 * | 19 | 47 713 422 | 29 713 | PC | EH domain-containing 2 | | | −1.75 | $7.0 \times 10^{-7}$ |
| FSD1L** | 9 | 105 447 796 | 104 638 | PC | Fibronectin type III and SPRY domain-containing 1-like | 0.57 | $2.0 \times 10^{-6}$ | 0.75 | $7.7 \times 10^{-4}$ |

* Not aligned in STAR.
** Counts not significant in salmon.

**Table S4**. Transcriptome-wide significant genes for predicting ALS-FRS-R lower limb subscores with log2 fold change and Benjamini-Hochberg adjusted p-values reported for STAR and Salmon. Positive fold change relates to better prognostic scoring. PC: protein coding; PP: processed pseudogene; UP: unprocessed pseudogene, snoRNA: small nucleolar RNA; TUP: translated unprocessed pseudogene; lncRNA: long non-coding RNA.

| | | | | | | STAR | | | Salmon | | |
|---|---|---|---|---|---|---|---|---|---|---|---|
| Name | Chr | Start | Length | Type | Description | $\log_2$ FC | $\log_2$ SE | $p$ | $\log_2$ FC | $\log_2$ SE | $p$ |
| AC103925.1 | 1 | 201 222 113 | 1011 | lncRNA | Novel transcript | 0.54 | 0.13 | $2.7 \times 10^{-5}$ | — | — | — |
| AC138866.1 | 5 | 70 219 918 | 39 013 | UP | Gusb pseudogene 14 | −0.12 | 0.03 | $6.8 \times 10^{-6}$ | −0.15 | 0.04 | $1.1 \times 10^{-4}$ |
| AC138866.2 | 5 | 70 197 255 | 10 491 | UP | Glucuronidase, beta (gusb) pseudogene | −0.12 | 0.03 | $8.3 \times 10^{-6}$ | 0.12 | 0.11 | $2.8 \times 10^{-1}$ |
| AL359955.2 | 9 | 64 737 403 | 353 | PP | Pseudogene similar to tripartite motif-containing protein | −0.26 | 0.05 | $4.0 \times 10^{-7}$ | — | — | — |
| AL591379.2 | 9 | 63 025 597 | 397 | UP | Pseudogene similar to part of tripartite motif-containing 43 (trim43) | −0.24 | 0.05 | $4.8 \times 10^{-7}$ | — | — | — |
| AL732366.2 | X | 20 136 066 | 70 | snoRNA | — | −0.24 | 0.05 | $9.5 \times 10^{-6}$ | −0.20 | 0.07 | $3.8 \times 10^{-3}$ |
| ARHGAP45 | 19 | 1 065 923 | 20 706 | PC | Rho gtpase activating protein 45 | 0.03 | 0.04 | $4.0 \times 10^{-1}$ | 0.32 | 0.06 | $1.2 \times 10^{-7}$ |
| BX284632.2 | 9 | 65 798 726 | 399 | PP | Pseudogene similar to part of tripartite motif-containing protein family | −0.24 | 0.05 | $4.8 \times 10^{-7}$ | — | — | — |
| CPEB1 | 15 | 82 543 201 | 105 661 | PC | Cytoplasmic polyadenylation element binding protein 1 | 0.19 | 0.06 | $2.7 \times 10^{-3}$ | 0.39 | 0.08 | $2.8 \times 10^{-6}$ |
| FOSB | 19 | 45 467 995 | 7185 | PC | Fosb proto-oncogene, ap-1 transcription factor subunit | −0.70 | 0.17 | $2.9 \times 10^{-5}$ | −0.40 | 0.12 | $6.4 \times 10^{-4}$ |
| GUSBP1 | 5 | 21 341 833 | 247 540 | TUP | Gusb pseudogene 1 | −0.12 | 0.03 | $9.0 \times 10^{-6}$ | −0.14 | 0.06 | $1.6 \times 10^{-2}$ |
| GUSBP2 | 6 | 26 871 484 | 85 071 | TUP | Gusb pseudogene 2 | −0.11 | 0.02 | $1.0 \times 10^{-5}$ | −0.02 | 0.03 | $3.9 \times 10^{-1}$ |
| GUSBP3 | 5 | 69 639 459 | 39 801 | TUP | Gusb pseudogene 3 | −0.11 | 0.03 | $1.9 \times 10^{-5}$ | −0.05 | 0.03 | $1.4 \times 10^{-1}$ |
| **IGFN1** | 1 | 201 190 825 | 38 128 | PC | Immunoglobulin like and fibronectin type iii domain containing 1 | 0.79 | 0.14 | $4.2 \times 10^{-8}$ | 0.80 | 0.15 | $8.0 \times 10^{-8}$ |
| IL18R1 | 2 | 102 311 529 | 87 247 | PC | Interleukin 18 receptor 1 | 0.08 | 0.06 | $1.5 \times 10^{-1}$ | 0.40 | 0.09 | $3.1 \times 10^{-6}$ |
| MDC1 | 6 | 30 699 807 | 17 641 | PC | Mediator of dna damage checkpoint 1 | −0.08 | 0.02 | $8.1 \times 10^{-6}$ | −0.01 | 0.07 | $8.4 \times 10^{-1}$ |
| **PBX1** | 1 | 164 555 584 | 343 713 | PC | Pbx homeobox 1 | −0.10 | 0.02 | $7.5 \times 10^{-7}$ | −0.15 | 0.03 | $3.8 \times 10^{-6}$ |
| PICART1 | 17 | 50 050 349 | 5391 | lncRNA | P53 inducible cancer associated rna transcript 1 | −0.27 | 0.06 | $1.7 \times 10^{-5}$ | −0.31 | 0.07 | $2.5 \times 10^{-5}$ |
| SLC16A3 | 17 | 82 228 397 | 32 733 | PC | Solute carrier family 16 member 3 | 0.34 | 0.08 | $2.8 \times 10^{-5}$ | 0.33 | 0.08 | $1.2 \times 10^{-4}$ |
| **TMEM9** | 1 | 201 134 772 | 36 803 | PC | Transmembrane protein 9 | 0.16 | 0.03 | $5.0 \times 10^{-8}$ | 0.16 | 0.03 | $3.7 \times 10^{-6}$ |

**Table S5**. Functional consequences of significant isoform switches annotated by the number of genes falling under it, as well as directionality if any for that feature. SAA2 is bolded as it is also transcriptome-wide significant.

| Mechanism | | Consequence | | |
|---|---|---|---|---|
| Feature | $n$ | Direction | $n$ | Genes |
| Genomic domain position | 16 | | 16 | ARHGEF6 CDKN1A MAGED2 MPST MYH3 ORAI1 PIP5K1B PLCE1 RUFY1 **SAA2** SAC3D1 SAMD4A SCHIP1 TP53INP1 TPST2 ZC3H6 |
| Isoform length | 14 | Gain | 8 | CDKN1A MPST MYH3 ORAI1 PIP5K1B PLCE1 SAMD4A TPST2 |
| | | Loss | 6 | ARHGEF6 FAM78A RUFY1 **SAA2** SAC3D1 SCHIP1 |
| Intron structure | 13 | | 13 | ARHGEF6 CDKN1A FAM78A MAGED2 MPST PLCE1 RUFY1 **SAA2** SAMD4A SCHIP1 TP53INP1 TPST2 ZC3H6 |
| TSS | 13 | Upstream | 7 | MAGED2 MYH3 PIP5K1B PLCE1 SAMD4A SCHIP1 TPST2 |
| | | Downstream | 6 | ARHGEF6 CDKN1A FAM78A MPST RUFY1 SAC3D1 |
| 5' UTR seq similarity | 12 | Longer | 7 | ARHGEF6 CDKN1A FAM78A MPST ORAI1 PIP5K1B RUFY1 |
| | | Shorter | 5 | MAGED2 MYH3 SAC3D1 SCHIP1 TPST2 |
| ORF genomic | 12 | | 12 | ARHGEF6 FAM78A MPST MYH3 ORAI1 PIP5K1B PLCE1 RUFY1 **SAA2** SAMD4A SCHIP1 TP53INP1 |
| 5' UTR length | 10 | Longer | 6 | ARHGEF6 CDKN1A MPST ORAI1 PIP5K1B RUFY1 |
| | | Shorter | 4 | MYH3 SAC3D1 SCHIP1 TPST2 |
| Isoform seq similarity | 10 | Longer | 7 | MPST MYH3 ORAI1 PIP5K1B PLCE1 SAMD4A TPST2 |
| | | Shorter | 3 | FAM78A RUFY1 **SAA2** |
| ORF length | 10 | Shorter | 6 | ARHGEF6 FAM78A MPST ORAI1 RUFY1 TP53INP1 |
| | | Full gain | 2 | PLCE1 SAMD4A |
| | | Longer | 2 | MYH3 PIP5K1B |
| Exon number | 9 | Gain | 6 | FAM78A MYH3 PIP5K1B PLCE1 SAMD4A TP53INP1 |
| | | Loss | 3 | ARHGEF6 ORAI1 RUFY1 |
| IDR identified | 9 | IDR loss | 6 | ARHGEF6 ORAI1 PIP5K1B RUFY1 SCHIP1 TP53INP1 |
| | | IDR gain | 3 | MYH3 PLCE1 SAMD4A |
| ORF seq similarity | 9 | Shorter | 5 | ARHGEF6 FAM78A ORAI1 RUFY1 TP53INP1 |
| | | Full gain | 2 | PLCE1 SAMD4A |
| | | Longer | 2 | MYH3 PIP5K1B |
| TTS | 9 | Downstream | 5 | PIP5K1B PLCE1 **SAA2** SAMD4A TPST2 |
| | | Upstream | 4 | FAM78A MYH3 ORAI1 RUFY1 |
| 3' UTR length | 8 | Longer | 5 | MYH3 ORAI1 PIP5K1B TP53INP1 TPST2 |
| | | Shorter | 3 | FAM78A RUFY1 **SAA2** |
| 3' UTR seq similarity | 7 | Longer | 4 | MYH3 ORAI1 PIP5K1B TPST2 |
| | | Shorter | 3 | FAM78A RUFY1 **SAA2** |
| Domains identified | 6 | Gain | 4 | MYH3 PIP5K1B PLCE1 SAMD4A |
| | | Loss | 2 | ARHGEF6 RUFY1 |
| Last exon | 5 | Downstream | 3 | MYH3 PIP5K1B PLCE1 |
| | | Upstream | 2 | RUFY1 **SAA2** |
| Coding potential | 3 | Coding | 3 | PLCE1 **SAA2** SAMD4A |
| Intron retention | 3 | Loss | 2 | MPST SAC3D1 |
| | | Gain | 1 | ORAI1 |
| Domain length | 1 | Gain | 1 | MYH3 |
